# Whole exome sequencing and replication for breast cancer among Hispanic/Latino women identifies *FANCM* as a susceptibility gene for estrogen-receptor-negative breast cancer

**DOI:** 10.1101/2023.01.25.23284924

**Authors:** Jovia L. Nierenberg, Aaron W. Adamson, Donglei Hu, Scott Huntsman, Carmina Patrick, Min Li, Linda Steele, Barry Tong, Yiwey Shieh, Laura Fejerman, Stephen B. Gruber, Christopher A. Haiman, Esther M. John, Lawrence H. Kushi, Gabriela Torres-Mejía, Charité Ricker, Jeffrey N. Weitzel, Elad Ziv, Susan L. Neuhausen

## Abstract

**Introduction:** Breast cancer (BC) is one of the most common cancers globally. Genetic testing can facilitate screening and risk-reducing recommendations, and inform use of targeted treatments. However, genes included in testing panels are from studies of European-ancestry participants. We sequenced Hispanic/Latina (H/L) women to identify BC susceptibility genes.

**Methods:** We conducted a pooled BC case-control analysis in H/L women from the San Francisco Bay area, Los Angeles County, and Mexico (4,178 cases and 4,344 controls). Whole exome sequencing was conducted on 1,043 cases and 1,188 controls and a targeted 857-gene panel on the remaining samples. Using ancestry-adjusted SKAT-O analyses, we tested the association of loss of function (LoF) variants with overall, estrogen receptor (ER)-positive, and ER-negative BC risk. We calculated odds ratios (OR) for BC using ancestry-adjusted logistic regression models. We also tested the association of single variants with BC risk.

**Results:** We saw a strong association of LoF variants in *FANCM* with ER-negative BC (p=4.1×10^−7^, OR [CI]: 6.7 [2.9-15.6]) and a nominal association with overall BC risk. Among known susceptibility genes, *BRCA1* (p=2.3×10^−10^, OR [CI]: 24.9 [6.1-102.5]), *BRCA2* (p=8.4×10^−10^, OR [CI]: 7.0 [3.5-14.0]), and *PALB2* (p=1.8×10^−8^, OR [CI]: 6.5 [3.2-13.1]) were strongly associated with BC. There were nominally significant associations with *CHEK2, RAD51D*, and *TP53*.

**Conclusion:** In H/L women, LoF variants in *FANCM* were strongly associated with ER-negative breast cancer risk. It previously was proposed as a possible susceptibility gene for ER-negative BC, but is not routinely tested in clinical practice. Our results demonstrate that *FANCM* should be added to BC gene panels.

## INTRODUCTION

Breast cancer is the most common cancer in women,^1^ and is influenced by hormonal, environmental, and genetic factors.^2^ Approximately 11% of screening age women have a first-degree relative diagnosed with breast cancer,^3^ and these women have ∼2-fold higher risk of being diagnosed with breast cancer.4 Germline pathogenic variants found in high-penetrance genes such as *BRCA1*^5^, *BRCA2*,^6^ *PALB2*,^7^ *TP53*^8,9^ and others^10^ are associated with high risk of breast cancer and underlie familial cancer syndromes. Several intermediate-penetrance genes including *CHEK2*^11^ and *ATM*^12^ also have been identified. In addition, genome-wide association studies (GWAS) have identified many common variants that contribute to breast cancer risk.^13^ However, all the pathogenic variants in susceptibility genes and the common breast cancer risk variants identified to date explain only half of the heritability of the disease.^13^

Most of the knowledge of genetic susceptibility to breast cancer is based on studies done in European-ancestry populations, leaving gaps in the understanding of genetic effects in other populations. In particular, among Hispanic/Latina (H/L) women in the United States (US), breast cancer is the most common cancer and leading cause of cancer-related death.^14^ Latin-American populations are genetically diverse, and many H/L individuals have admixed ancestry, comprised primarily of European, Indigenous American, and African components. ^15,16^ GWAS of breast cancer in H/L women led to the discovery of a protective variant near *ESR1*, which most commonly occurs in women with higher Indigenous American ancestry.^17^ In addition, unique founder variants have been identified in *BRCA1*,^18^ *PALB2*,^19,20^ and *CHEK2*.^19^ Since the sample sizes in studies of H/L women are smaller than in studies of White women in the U.S. and European women, the genetic contribution to breast cancer in this population remains poorly understood.

Genetic testing for pathogenic variants in breast cancer susceptibility genes is currently used to identify women at high risk of developing breast cancer, who may benefit from increased screening and risk-reducing interventions, for cascade testing in families to identify other individuals at increased risk, and to inform the use of targeted treatments in those who develop cancer.^21–24^ However, most evidence supporting association with risk in these genes is from studies of European ancestry participants. Two large recent studies in predominantly European ancestry participants confirm the association of known breast cancer susceptibility genes with increased breast cancer risk in population-based cohorts, reiterating that many of these genes are important to include on clinical genetic testing panels.^25,26^

To better understand the impact of rare variants in coding sequence of genes on breast cancer risk among H/L women, we performed a whole exome sequencing (WES) and targeted replication approach in over 8,500 H/L women from California and Mexico. We report an exome-wide significant association between *FANCM* and estrogen-receptor (ER) negative breast cancer, as well as associations between several other known risk genes and breast cancer.

## Methods

### Study samples

Our study was a discovery and replication pooled case-control analysis of invasive breast cancer among self-identified H/L women. All cases had been diagnosed with at least one invasive breast cancer and we used age at first breast cancer diagnosis for women diagnosed with more than one breast cancer at different ages. Participant selection in our discovery population has been previously described.^19^ Briefly, discovery cases were selected for having previously tested negative for *BRCA1/2*, and were diagnosed at <51 years of age and/or had bilateral (synchronous or metachronous) breast cancer, breast and ovarian cancers, or were diagnosed between 51 and 70 years with a family history of breast cancer in ≥ 2 first-degree or second-degree relatives diagnosed at age <70 years. Discovery cases were selected from three high-risk registry studies. We included self-identified H/L women with breast cancer from the Clinical Cancer Genomics Community Research Network (CCGCRN),^27,28^ a network of cancer centers and community-based clinics that provide genetic counseling to individuals with a personal or family history of cancer. We also included self-identified H/L women with breast cancer from the University of California at San Francisco (UCSF) Clinical Genetics and Prevention Program and the University of Southern California (USC) Norris Comprehensive Cancer Center clinical genetics program. Discovery controls were self-identified H/L women enrolled by City of Hope (COH) staff through health fairs and participants in the Multiethnic Cohort (MEC). The MEC is a large prospective cohort study conducted in California (mainly Los Angeles county) and Hawaii.^29^ Controls from the MEC did not have breast cancer and approximately half had diabetes.

Self-identified H/L participants in the replication set were from six studies (**Table 1**). The Cancer de Mama (CAMA) study is a population-based case–control study of breast cancer conducted in Mexico City, Monterrey and Veracruz. Cases aged 35–69 years at diagnosis and diagnosed between 2004 and 2007 were recruited from 12 hospitals (3 to 5 hospitals in each region). Controls were recruited based on membership in the same health plan as the cases and were frequency-matched on 5-year age groups.^30,31^ The California Pacific Medical Center - Breast Health Center (CPMC) cohort^32^ is composed of women who presented for mammography in San Francisco, California between 2004 and 2011 and we included incident and prevalent breast cancer cases. The PATHWAYS study is a cohort of breast cancer cases diagnosed at Kaiser Permanente Northern California.^33^ We used samples from participants who were enrolled in PATHWAYS and reported H/L ethnicity. From the nested case-control study within the MEC, we included cases with invasive breast cancer diagnosed at the age of >50 years and controls matched on age and self-identified ethnicity.^29^ The Northern California Breast Cancer Family Registry (NC-BCFR)^34^ recruited and followed about 4,000 breast cancer families and individuals with breast cancer, including cases with indicators of increased genetic susceptibility (including diagnosis before age 35, personal history of ovarian cancer, personal history of first breast cancer in contralateral breast before age 50, family history of breast cancer in first degree relative, family history of ovarian cancer in first degree relative, or family history of childhood cancer cancer in first degree relative) and cases without such indicators. We included both subsets of H/L cases from the NC-BCFR. Cases aged 18–64 years diagnosed from 1995 to 2009 were ascertained through the Greater Bay Area Cancer Registry. Population controls were identified through random-digit dialing and frequency matched on race/ethnicity and 5-year age groups to cases diagnosed from 1995 to 1998. The San Francisco Bay Area Breast Cancer Study (SFBCS)^35^ is a population-based multiethnic case–control study of breast cancer, where cases aged 35–79 years at diagnosis with invasive breast cancer from 1995 to 2002 were identified through the Greater Bay Area Cancer Registry and controls were identified by random-digit dialing and matched to cases on race/ethnicity and 5-year age groups. All participants were consented and enrolled into the study through center-specific institutional review board-approved protocols.

**Table 1:**
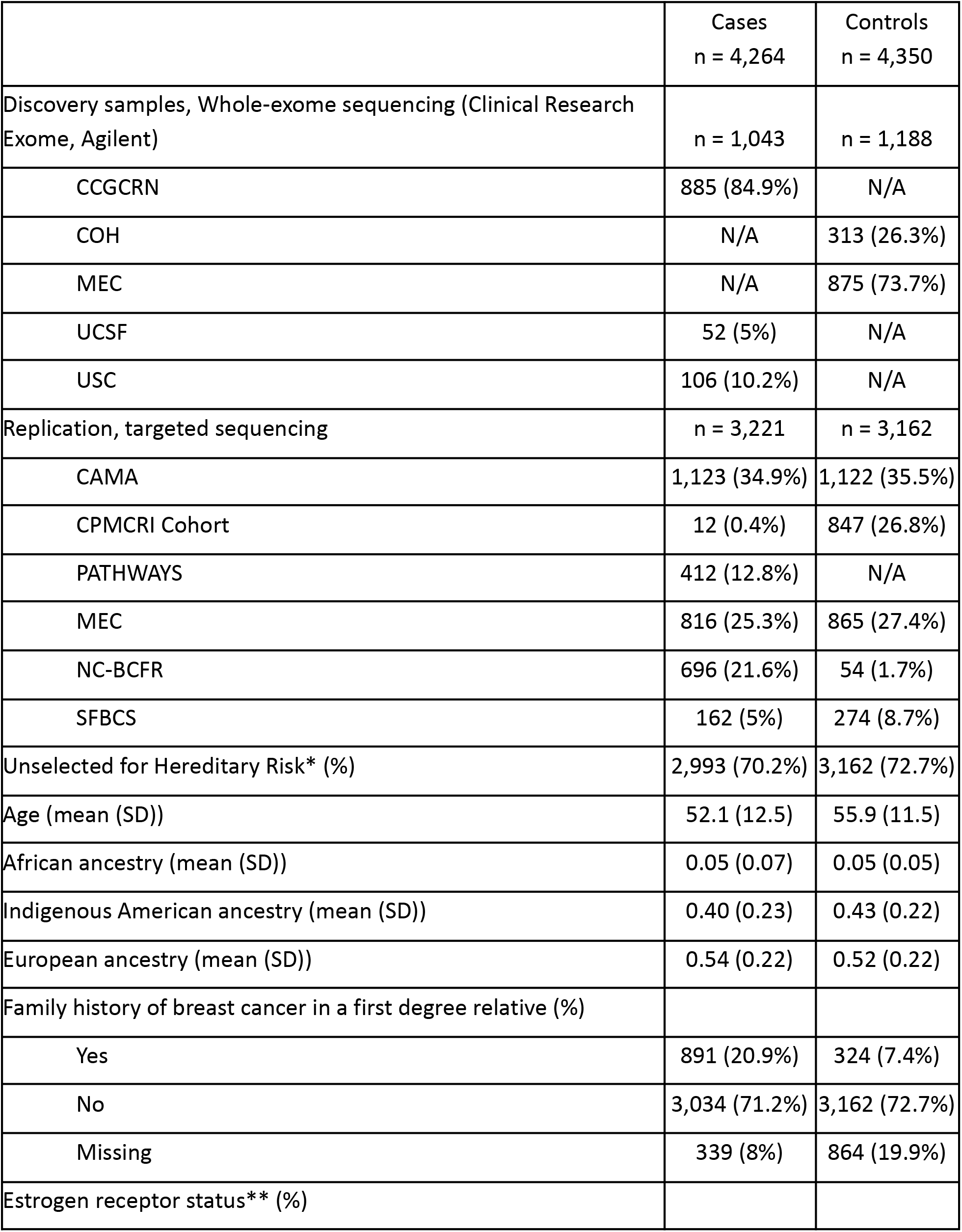
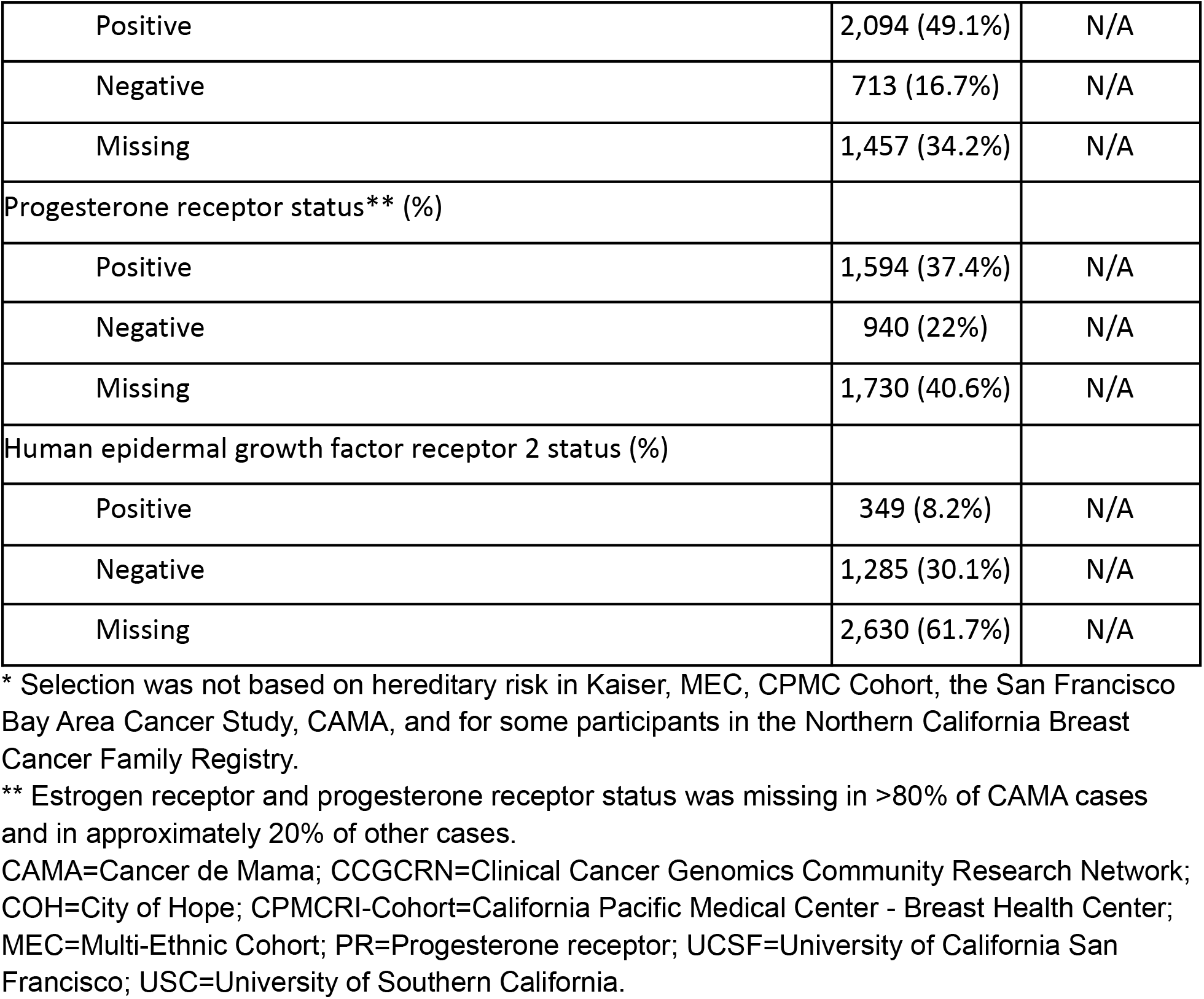
Characteristics of Study Participants.

### Sequencing and genotyping

Whole exome sequencing (WES) from DNA of discovery participants has been described previously.^19^ The SureSelect Clinical Research Exome (Agilent, Santa Clara, CA) kit was used to capture exons of all known human transcripts. For participants in the replication set, targeted sequencing was conducted on 857 genes, which were selected based on the discovery data and biological plausibility. A custom SureSelect XT kit (Agilent, Santa Clara, CA) was used to capture the exons of 857 genes and also included 189 known breast cancer SNPs and 100 ancestry informative markers (Supplemental Table X). Briefly, for both the WES and the targeted sequencing, we used KAPA Hyper Preparation Kits (Kapa Biosystems, Inc., Wilmington, MA) to generate libraries from 500 ng DNA. One hundred base-pair paired end sequencing on the HiSEQ 2500 Genetic Analyzer (Illumina Inc., San Diego, CA) was performed in the COH Integrative Genomics Core (IGC) to an average fold coverage of ×65 for the WES samples and ∼x75 for the targeted-sequencing samples. Paired-end reads from each sample were aligned to human reference genome (hg37) using the Burrows-Wheeler Alignment Tool (BWA, version 0.7.5a-r405) under default settings, and the aligned binary format sequence (BAM) files were sorted and indexed using SAMtools.^36,37^ The same FASTA reference file had been used for aligning the MEC control samples. The sorted and indexed BAMs were processed by Picard MarkDuplicates (version 1.67, http://broadinstitute.github.io/picard/) to remove duplicate sequencing reads.

Variant calling from the BAM files from the IGC and the Broad Sequencing Center were processed together at UCSF. After local realignment of reads around in-frame insertions and deletions (indels) and base quality score recalibration by The Genome Analysis Toolkit (GATK, v3.6-0-g89b7209), GATK HaplotypeCaller was used to call variants (https://software.broadinstitute.org/gatk). Variants with a call quality <20, a read depth <10, a less frequent allele depth of <4, or an allele fraction ratio <30% were filtered out for low quality. We also excluded participants with <20-fold average coverage. DNA from eight MEC participants were sequenced at both COH and the Broad Sequencing Center with >99.8% concordance for variant calling.

After sequencing, we excluded discovery cases with previously undetected *BRCA1/2* pathogenic variants. We used PLINK 1.9 (http://www.cog-genomics.org/plink/1.9/)^38^ to exclude first-degree relatives within the discovery and replication separately and genetic duplicates across the discovery and replication. Due to differences in participant inclusion criteria among studies in our discovery sample, we conducted a control-control analysis and excluded variants that were different between our two control populations at an alpha threshold of 0.05. We also conducted a sensitivity analysis among participants of the CCGCRN/COH only, and excluded variants where the sensitivity analysis point estimate was outside of the 95% confidence interval of the main analysis.

### Ancestry estimation

Ancestry estimation for our discovery population has been described previously.^19^ The Clinical Research Exome included a custom panel of 180 ancestry-informative single nucleotide polymorphisms. On the basis of a previous publication,^39^ these markers were selected to be informative for ancestry in mixed-European, Indigenous-American, and African populations. In addition, we selected 7,691 variants common to our WES data and a data set of Axiom arrays, including African (N = 90), European (N= 90), and Indigenous American (N = 51) populations. We selected unlinked markers by linkage disequilibrium pruning in PLINK, identifying a subset of 4,544 variants for ancestry estimation. We estimated genetic ancestry using ADMIXTURE 1.3 ^40^ and performed analyses with both supervised (specifying the ancestral populations) and unsupervised (including the data from ancestral populations, but not specifying the identity of ancestral populations) runs. To determine genetic ancestry in the MEC control participants, we used the same ancestral reference samples and the same approach using ADMIXTURE. However, since the exome sequencing data did not include ancestry informative markers, we selected a subset of independent variants (n = 12,758) that overlapped between the Axiom arrays and the MEC dataset.

In our replication sample, we performed genetic ancestry estimation for each individual. We used 90 European (1000 Genomes), 90 African (1000 Genomes), 90 east Asian (1000 Genomes) and 71 Indigenous American ancestry^41^ reference samples. We combined our study data and the reference data using 1,195 SNPs that overlapped across all datasets. We then used ADMIXTURE 1.3 to estimate the ancestry for each individual using the supervised method.

### Statistical analysis

Gene-based aggregate rare variant analyses were based on loss of function (LoF) variants, including frameshift, stopgain, and splice variants. Variants with minor allele frequency > 0.0025 and benign clinical significance in CLINVAR^42^ were excluded from the gene-based analyses. Statistical significance for each gene was determined using SKAT-O.^43,44^ Odds ratios (OR) and 95% confidence intervals (CI) for breast cancer associated with any LoF variant for each gene were calculated using logistic regression models in which women with at least one LoF variant in a gene were encoded as “1” and all other women were encoded as “0”. These models included ancestry (European and Indigenous American) as covariates. These models constituted our burden tests. For additional quality control, results from SKAT-O and burden tests were included if the gene had > 5 variants or at least one alternate allele in both cases and controls. We used the replication sample for all *BRCA1/2* analyses, as *BRCA1/2* status was a selection criterion for the discovery sample. Additionally, we looked for the known Mexican founder *BRCA1* exon 9-12 deletion, and verified its presence using manual review with the Integrative Genomics Viewer (IVG), then included this mutation in the exome data. Single variant analyses and aggregate rare variant analyses were conducted in discovery and replication analyses separately, as well as in a joint discovery and replication analysis. Additional breast cancer subtype-specific analyses were conducted, where cases were separately restricted to ER-positive and ER-negative disease. Sensitivity analyses were run separately in participants from hereditary breast cancer studies and studies that did not select cases based on breast cancer risk factors. All analyses were adjusted for European and Indigenous American ancestry.

## Results

The mean age at diagnosis of breast cancer cases was 42.6 years (standard deviation [SD]: 8.5) and the mean age at enrollment of controls was 60.3 years (SD: 10.9) in the discovery set (P=2.4×10^−200^) (**Table 1**). In the replication set, the mean age at diagnosis of breast cancer was 55.3 years (SD: 12.0) and the mean age of controls at enrollment was 54.9 years (SD: 11.4, P=0.27). European ancestry (EA) proportion was slightly higher among cases (P=6.0×10^−6^) and Indigenous American ancestry (IA) was slightly higher among controls (P=4.4×10^−7^). Ancestry proportions for cases and controls are shown in **Supplementary Figure 1**. History of a first-degree relative with breast cancer was higher among cases than controls (P=2.5×10^−108^). Most of the cases were ER-positive and progesterone receptor (PR)-positive and 21.3% of those who were tested were human epidermal growth factor receptor 2 (HER2)-positive.

We conducted joint analyses of the discovery and replication datasets. In the analysis that tested for overall breast cancer risk, we found significant associations with *BRCA1, BRCA2* and *PALB2*, with ORs (95% CI) of 24.9 (6.1-102.5), 7.0 (3.5-14.0), and 6.5 (3.2-13.1), respectively. In the analyses for ER-negative and ER-positive breast cancers, we found significant associations with ER-negative breast cancer for LoF variants in *FANCM, BRCA1*, and *BRCA2* with ORs of 6.7 (2.9-15.6), 40.7 (8.9-186.5), and 10.5 (4.5-24.7), respectively (**Table 2** and **Figure 1**). No exome-wide significant associations were found with ER-positive breast cancer. Three other known breast cancer genes had suggestive associations with breast cancer in our study sample: *CHEK2* with overall and ER-positive breast cancer, *RAD51D* with ER-negative breast cancer, and *TP53* with ER-negative breast cancer (**Supplementary Table 1** and **Figure 1**). We found no significant associations of *ATM, BARD1, CDH1, RAD51C*, or *RECQL* and breast cancer risk. We ran a sensitivity analysis for *ATM* using truncating variants only, due to the unexpectedly null results and large number of splice variants found in this gene, and also found no association with breast cancer (OR=1.2 [0.8-1.9]). All genes with suggestive (P<0.01) associations for overall, ER-positive, and ER-negative breast cancer are shown in **Supplementary Tables 2** and **3**.

**Table 2:** Gene-Based P-Values from Joint Analysis with Exome-Wide Significance, for Breast Cancer Overall, ER-Positive, and ER-Negative Disease.

| Gene | Chr | Overall | ER-Positive | ER-Negative |
| --- | --- | --- | --- | --- |
| *BRCA1* * | 17 | **$2.3 \times 10^{-10}$** | 0.03 | **$4.4 \times 10^{-16}$** |
| *BRCA2* * | 13 | **$8.4 \times 10^{-10}$** | $3.3 \times 10^{-4}$ | **$8.0 \times 10^{-15}$** |
| *FANCM* | 14 | $9.8 \times 10^{-3}$ | 0.11 | **$4.1 \times 10^{-7}$** |
| *PALB2* | 16 | **$1.8 \times 10^{-8}$** | $1.3 \times 10^{-5}$ | $5.9 \times 10^{-5}$ |

Chr=Chromosome; ER=estrogen receptor.
P-values are from gene-based SKAT-O analyses. Bold P-values indicate Bonferroni corrected statistical significance at an alpha threshold of $0.05/20,000=2.5 \times 10^{-6}$ .
\* Discovery participants were selected for being *BRCA1/2* negative (see methods), replication results are presented for *BRCA1/2*.

**Figure 1:**
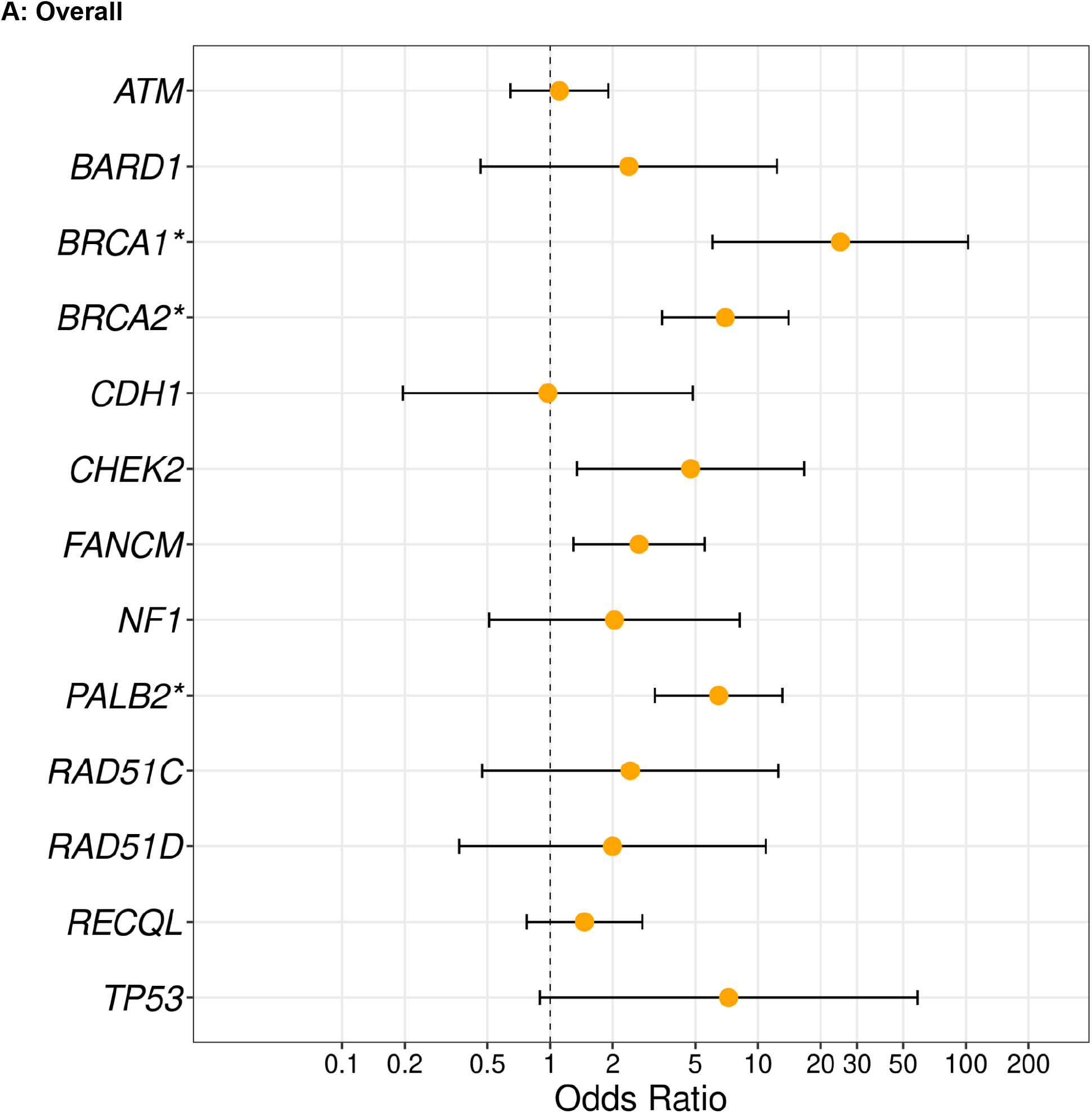
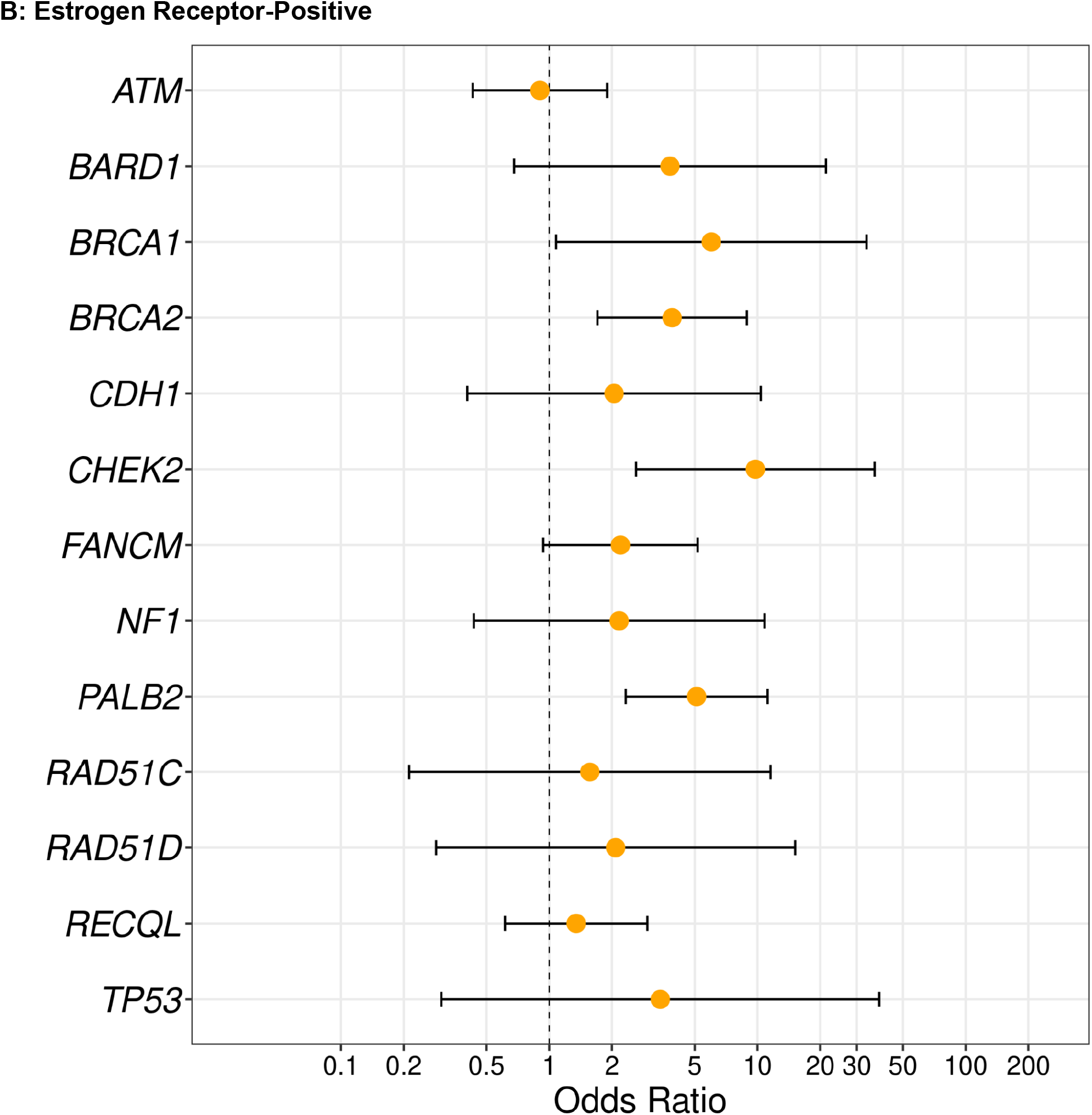
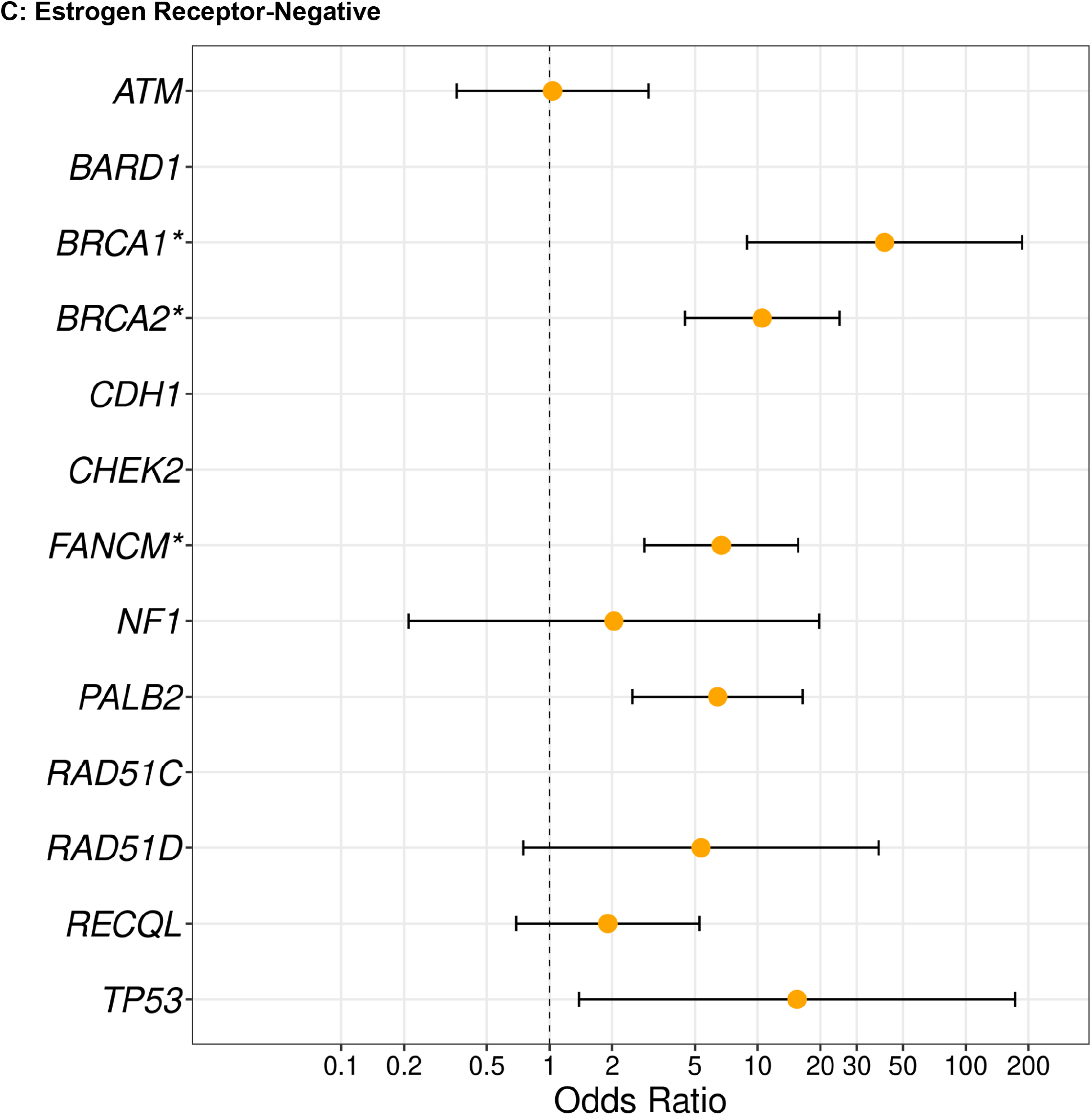
Gene-Based Odds Ratios from Joint Analysis for Breast Cancer Overall, ER-Positive, and ER-Negative Disease. Odds ratios and confidence intervals are presented for overall breast cancer (Panel A), estrogen receptor (ER)-positive (Panel B), and ER-negative (Panel C) disease. The orange dot represents the point estimate and the bars represent the upper and lower bound of the 95% confidence intervals. The X-axis describes the odds ratio on a log scale, the Y-axis represents the individual genes. Genes are listed in alphabetical order. Results were included for genes that had > 5 variants or at least one alternate allele in both cases and controls.

The association between *FANCM* LoF variants and ER-breast cancer was largely driven by two stopgain variants, rs147021911 (chr14:45658326C>T) and rs144567652 (chr14:45667921C>A / C>T) (**Figure 2**). A total of 1.7% of participants with ER-negative disease had a LoF variant in *FANCM*, compared to only 0.6% of cases with ER-positive breast cancer and 0.2% of controls (**Figure 2**). More than half of participants had a missense variant in *FANCM* and proportions were similar among participant groups (data not shown).

**Figure 2:**
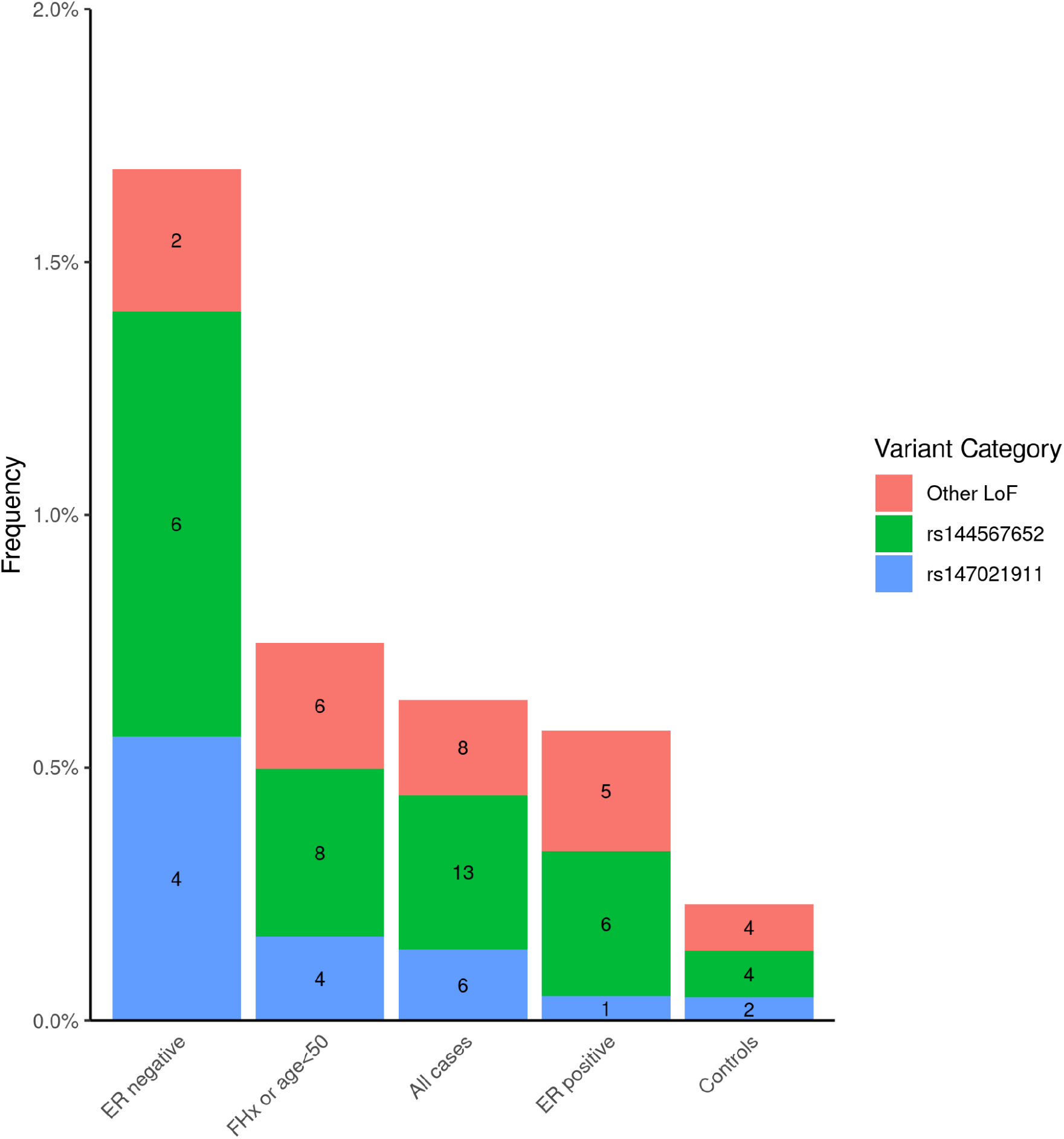
*FANCM* loss of function (LoF) Variant Frequencies by Participant Group. The X-axis shows groups of participants. The Y-axis shows the frequency of carriers (all carriers are heterozygous). The variant, rs144567652, is also known as chr14:45667921C>A / C>T, c.5791C>T, and p.Arg1931* (overall OR=3.2, 95% CI: 1.0-9.8 and ER-negative OR=8.1, 95% CI: 2.3-29.0). The variant, rs147021911, is also known as chr14:45658326C>T, c.5101 C >T, and p.Gln1701* (overall OR=3.0, 95% CI: 0.6-14.7 and ER-negative OR=11.3, 95% CI: 2.1-62.2). Other loss of function (LoF) variants found in FANCM include chr14:45605743G>A, rs140760056 (chr14:45633697C>T), rs368728266 (chr14:45636336C>T), rs778176467 (chr14:45642357C>T), chr14:45644584A>AT, rs1566762924 (chr14:45644795T>C), chr14:45644911, chr14:45644926TAG>T, rs755947203 (chr14:45653091C>A,T), rs930692973 (chr14:45658155C>G,T), and rs1379375089 (chr14:45665681C>T). ER=estrogen receptor; FHx=family history of breast cancer in first-degree relatives.

Since ascertainment can affect effect sizes and our discovery dataset and some of our replication datasets were selected based on several criteria, we analyzed the top genes among participants in the replication set who were from studies of cases unselected for hereditary risk (**Supplementary Figure 2**). The effect sizes for *CHEK2* and *PALB2* were similar in all studies and in unselected studies. The OR for *CHEK2* and ER-positive breast cancer in unselected studies was 10.7, which was similar to the OR of 10.8 found in all studies. We found that the association between *FANCM* and ER-negative breast cancer was similar in studies of unselected cases (OR=5.6) and in all studies (OR=6.7).

Since many of the known genes for breast cancer are involved in repair of double-stranded breaks, we reviewed all of the suggestive associations in the combined discovery and validation analyses and identified genes which are members of this pathway using a previously curated list.<u>^45^</u> In analyses that included both LoF variants and missense variants with high likelihood of being deleterious, we found suggestive evidence for *ATR* and *FANCG* (**Supplementary Tables 4** and **5**). Of these, *ATR* was more strongly associated with ER-positive disease, while *FANCG* was more strongly associated with ER-negative breast cancer (**Supplementary Table 4** and **5**).

## Discussion

In this case-control study of breast cancer in over 8,500 H/L women, we found strong evidence of association between breast cancer and LoF variants in *FANCM* largely driven by two *FANCM* stop gain variants. In addition we saw strong associations with the known breast cancer genes *BRCA1, BRCA2*, and *PALB2*.

The two most common *FANCM* variants we observed are more common in European populations than other world populations. These variants were both first identified in European ancestry *BRCA1/2*-negative familial breast cancer studies,^46,47^ and both have previously been associated with ER-negative or triple negative breast cancer.^46,48,49^ Additionally, rs147021911 (chr14:45658326C>T, c.5101 C >T, p.Gln1701*) was associated with poor breast cancer survival in a Finnish population,^50^ and other *FANCM* LoF mutations were found in small breast cancer case series,^51–55^ including a study that showed bi-allelic *FANCM* variants in early onset or bilateral breast cancer cases.^56^ To our knowledge, our study is the first to find an exome-wide significant association between *FANCM* and ER-negative breast cancer in any population. Since we used an exome-wide association approach for discovery, the effect size we detected may be inflated by the winner’ s curse.^57^ However, even if the effect size in our study is inflated due to these factors, the higher impact on ER-negative disease is consistent with prior studies and suggests that testing for *FANCM* LoF variants can help identify women at increased risk for developing ER-negative breast cancer.

Similar to most other breast cancer susceptibility genes, *FANCM* is involved in double-stranded DNA repair. In particular, FANCM localizes to stalled replication forks and initiates the response to double-stranded breaks by homologous recombination repair.^58^ *FANCM* is a member of the Fanconi Anemia (FA) complex and is the most conserved gene within the FA pathway, ^59^ although it has recently been excluded as a gene predisposing for FA.^56^ The FA pathway is essential for handling DNA interstrand cross linking damage in DNA repair through the homologous recombination repair pathway.^60^ A stronger association between *FANCM* and ER-negative disease than ER-positive disease is similar to results for *RAD51C* and *RAD51D*,^25^ which are also key components of the FA pathway.

*FANCM* previously has been proposed as a susceptibility gene for development of ER-negative breast cancer although without genome-wide significance,^25^ but is not routinely tested in clinical practice.^46^ Given our genome-wide significant result and the previous associations, the data now support inclusion of *FANCM* on clinical testing panels. Women with breast cancer and *FANCM* LoF variants may benefit from poly adenosine diphosphate-ribose polymerase (PARP) inhibitors;^49,61,62^ therefore, testing for FANCM LoF variants may identify additional therapeutic options for women with ER-negative breast cancer.

We found that carriers of LoF variants in *CHEK2* had a particularly high risk of ER-positive disease, though this did not reach exome-wide significance. The association with ER-positive disease among LoF variant carriers of *CHEK2* is consistent with prior studies;^25,26,63^ however, the effect sizes we saw in our study sample were substantially higher than those in previous reports.^25^ In analyses that included only cases that were not selected for familial breast cancer, the confidence intervals were wide but the lower bound of the confidence interval (2.1) included the OR found in the Breast Cancer Association Consortium study,^25^ indicating that our larger odds ratio may be due to chance. The higher OR we observed may also be due to the younger ages of the Latino population and/or to heterogeneity of genetic or hormonal and environmental risk factors across populations. Latinas tend to have more protective non-genetic risk factors such as younger age at first pregnancy, higher parity,^64^ lower postmenopausal hormone use, and lower alcohol consumption,^65^ when compared to US non-Hispanic White populations^66^ which may lead to stronger associations with genetic risk factors.^67^ In contrast to the strong association we observed with *CHEK2*, we found no evidence for an association with LoF variants in *ATM* in our study. The upper bound of the confidence interval for ER-positive disease excludes the reported odds ratio in the Breast Cancer Association Consortium study^25^ but not the odds ratio reported in the CARRIERS study.^26^ We have previously noted no significant association with *ATM* LoF variants in Latinas^19^ in a dataset that partially overlaps our current report. These results may be due to genetic or hormonal and environmental factors that attenuate the associations with *ATM* LoF variants in this population.

Our study, conducted among H/L women, identified strong associations with breast cancer risk genes, including the first exome-wide significant association between *FANCM* and ER-negative disease, with a notably smaller sample size than either of the recent rare exome variant association studies conducted in Europeans,^25,26^ thus demonstrating the importance of conducting genetic studies in admixed populations. Most large previous complex trait genetics studies, including those on breast cancer, have been conducted in European-ancestry participants.^68,69^ The lack of genetic studies in admixed populations exacerbates health disparities as genomics is increasingly used in clinical practice.

Our study has several strengths. To our knowledge, it is the largest exome sequencing analysis of breast cancer cases and controls in H/L women to date. We included participants from both hereditary-risk studies and unselected cases and controls, and compared results across these two study types, making our findings applicable to both groups. Our study also has several limitations. The WES for discovery was performed in only 1,043 cases and 1,188 controls and was likely underpowered to find intermediate-penetrance genes. To compensate for this, we selected 857 genes in the replication phase and sequenced these in 3,221 cases and 3,162 controls to enhance the likelihood of detecting associations. Larger studies in H/L populations are needed to confirm our results, to identify new candidate genes for breast cancer and to detect and understand factors contributing to heterogeneity of effect sizes compared to studies in US non-Hispanic White and European populations. Our discovery phase included sequencing data previously collected from a subset of controls in the Multi-Ethnic Cohort using a slightly different WES target capture. This difference potentially could lead to spurious associations due to technical or demographic differences between the different capture kits in the analysis of the discovery set. However, we accounted for potential demographic differences by adjusting for ancestry and excluding variants that were significantly different in an analysis of our two discovery control populations.

In conclusion, our study demonstrates an exome-wide significant association between LoF variants in *FANCM* and ER-negative breast cancer in H/L women from multiple studies. We also found exome-wide significant associations for *BRCA1, BRCA2*, and *PALB2*. Our findings suggest that *FANCM* should be added to genetic testing panels for breast cancer, which is especially important for H/L women. Additionally, our findings demonstrate the importance of conducting genetic studies in admixed populations.

## Data Availability

All data produced in the present study are available upon reasonable request to the authors

## Acknowledgements

We thank the participants of the SFBCS/NC-BCFR, Pathways, MEC, CAMA, and COH-CCGCRN studies.

This work was funded by the National Cancer Institute (R01CA184585, K24CA169004) and the State of California Initiative to Advance Precision Medicine (OPR18111). Research reported in this publication included work performed in the City of Hope Integrative Genomics Core supported by the National Cancer Institute of the National Institutes of Health under grant number P30CA033572. The content and views are solely the responsibility of the authors and should not be construed to represent the views of the National Institutes of Health. SLN and this research were partially funded by the Morris and Horowitz Families Professorship.

The Multiethnic Cohort was supported by NIH grants R01CA164973, R01CA054281, and CA063464. Sequencing of the MEC controls was conducted as part of the Slim Initiative for Genomic Medicine, a project funded by the Carlos Slim Health Institute in Mexico. Seventy-six of the controls were from the California Teachers Study; the collection and data were funded through the National Institutes of Health (R01CA77398). The Northern California Breast Cancer Family Registry was supported by grant UM1 CA164920 from the National Cancer Institute. The SFBCS was funded by grants R01CA063446 and R01CA077305 from the National Cancer Institute, grant DAMD17-96–1-6071 from the US Department of Defense, and grant 7PB-0068 from the California Breast Cancer Research Program. The Kaiser Permanente Research Program on Genes, Environment and Health was supported by Kaiser Permanente national and regional Community Benefit programs, and grants from the Ellison Medical Foundation, the Wayne and Gladys Valley Foundation, and the Robert Wood Johnson Foundation. The PATHWAYS cohort was supported by grants from the National Cancer Institute (U01 CA195565 R01 CA105274) The CAMA Study was funded by Consejo Nacional de Ciencia y Tecnologia, Mexico (SALUD-2002-C01–7462) and recruitment at the Puebla site was supported by R01CA120120.

LF is supported by R01CA204797. JNW was supported by NIH RC4 CA153828; Breast Cancer Research Foundation (#20-172), and American Society of Clinical Oncology Conquer Cancer® Research Professorship in Breast Cancer Disparities. YS was supported by the National Center for Advancing Translational Sciences under award KL2TR001870 and the National Cancer Institute under award K08CA237829.

## Tables and Figures

**Supplementary Figure 1:**
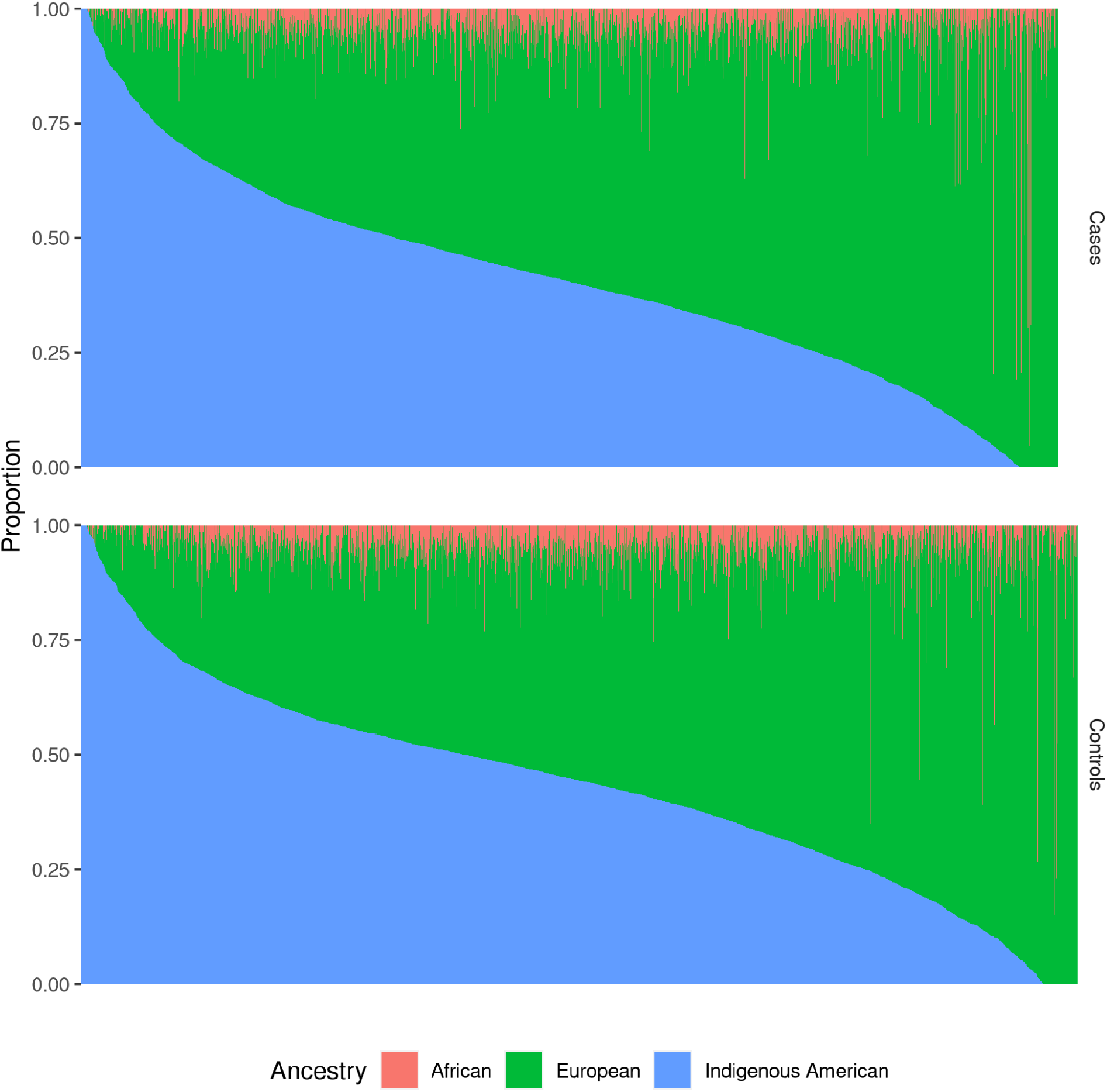
Ancestry Proportions among Cases and Controls. The Y-axis shows ancestry proportion, per individual, with all ancestry proportions adding up to 1.00. Each horizontal bar represents an individual. Individuals are sorted by proportion of indigenous ancestry from most on the left to least on the right. Ancestry was calculated using ADMIXTURE 1.3.

**Supplementary Figure 2:**
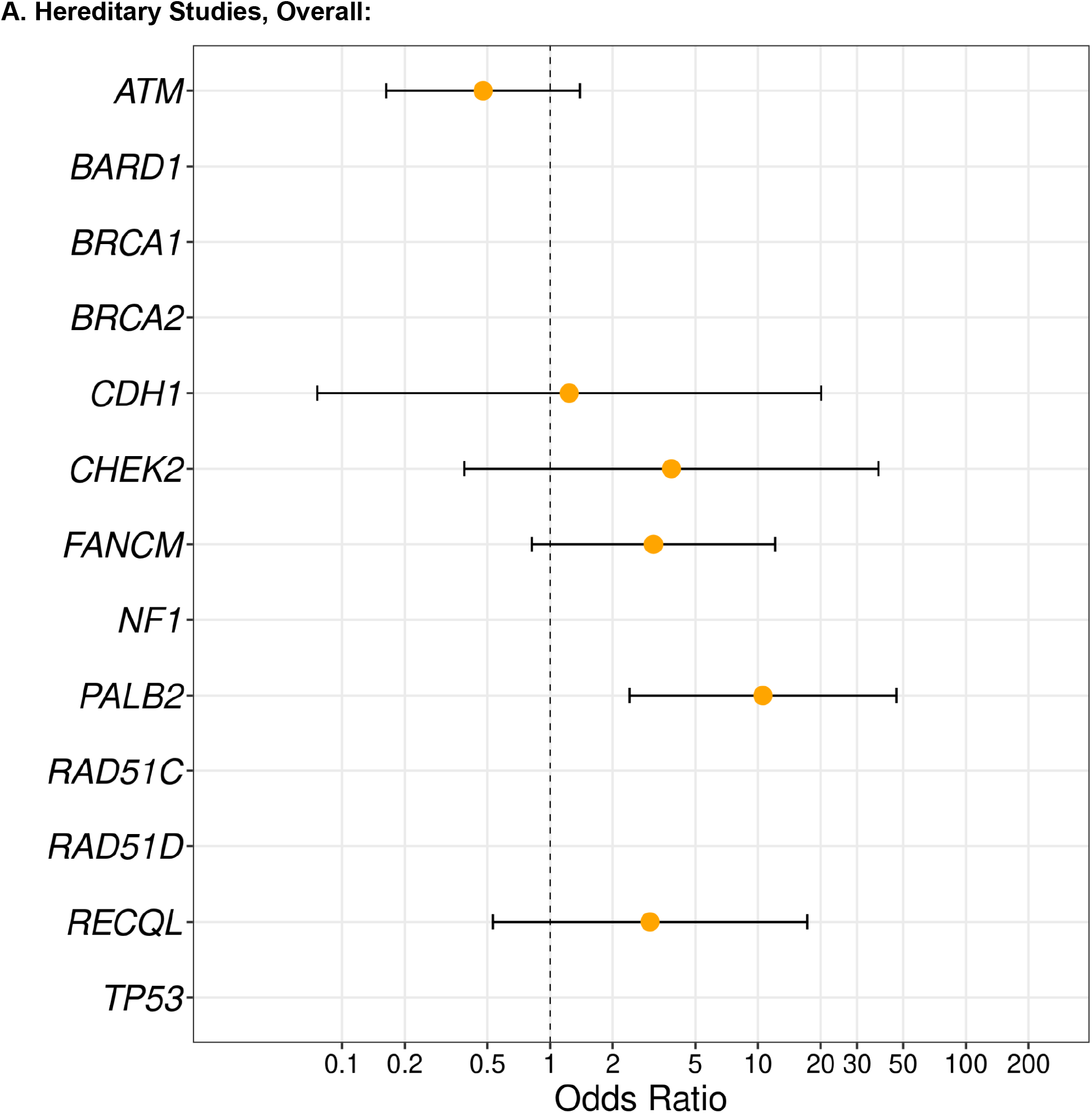
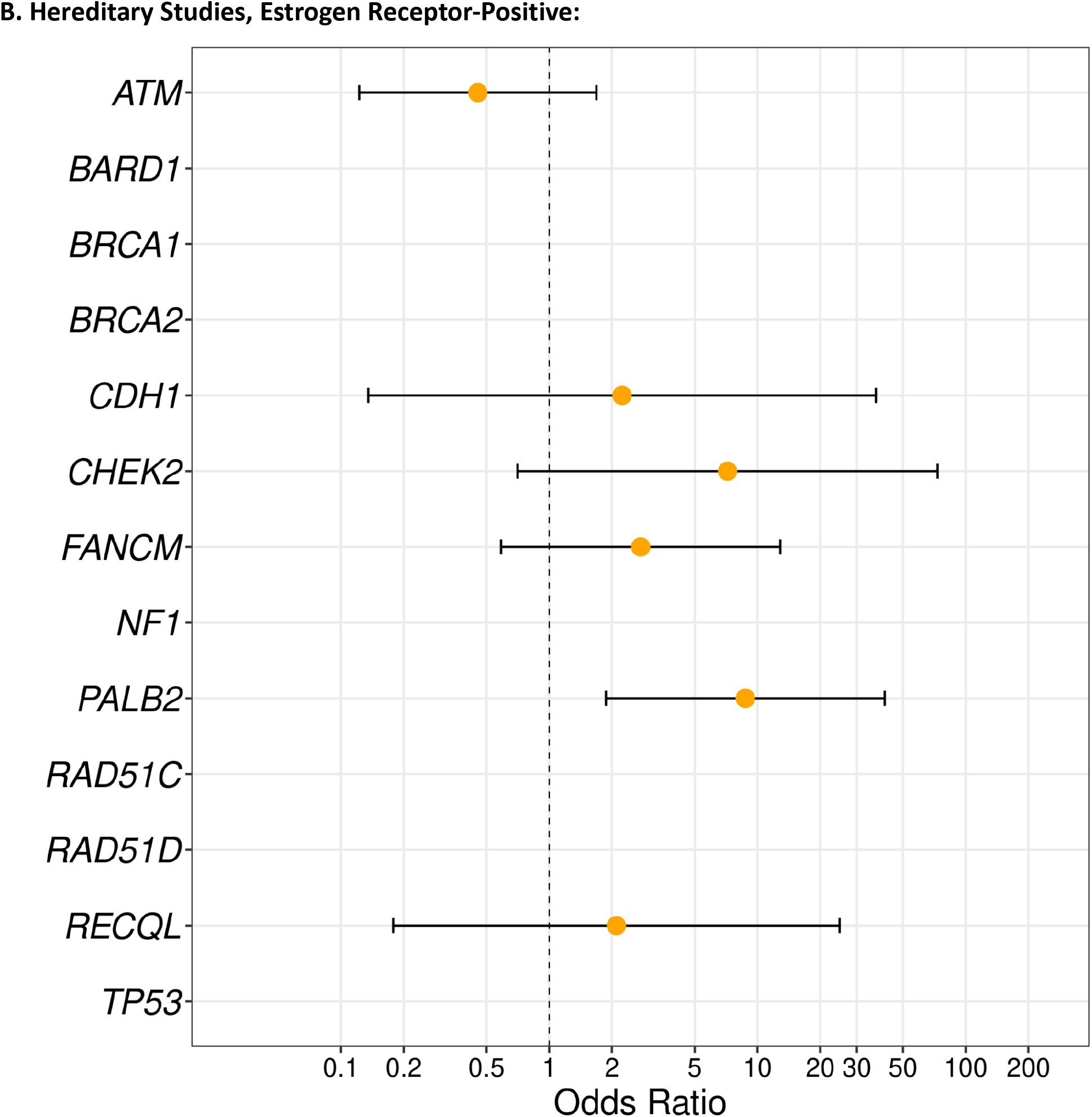
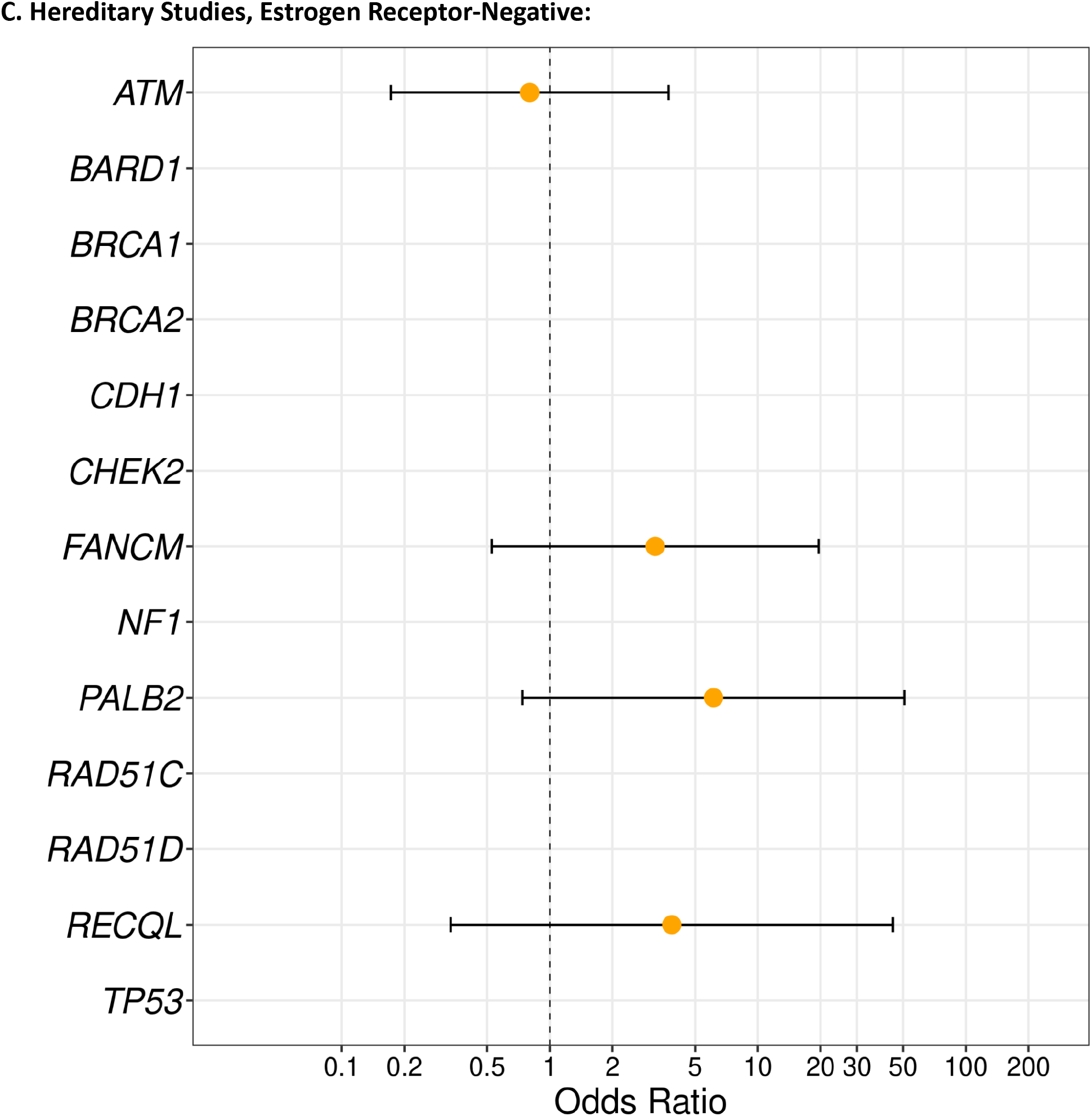
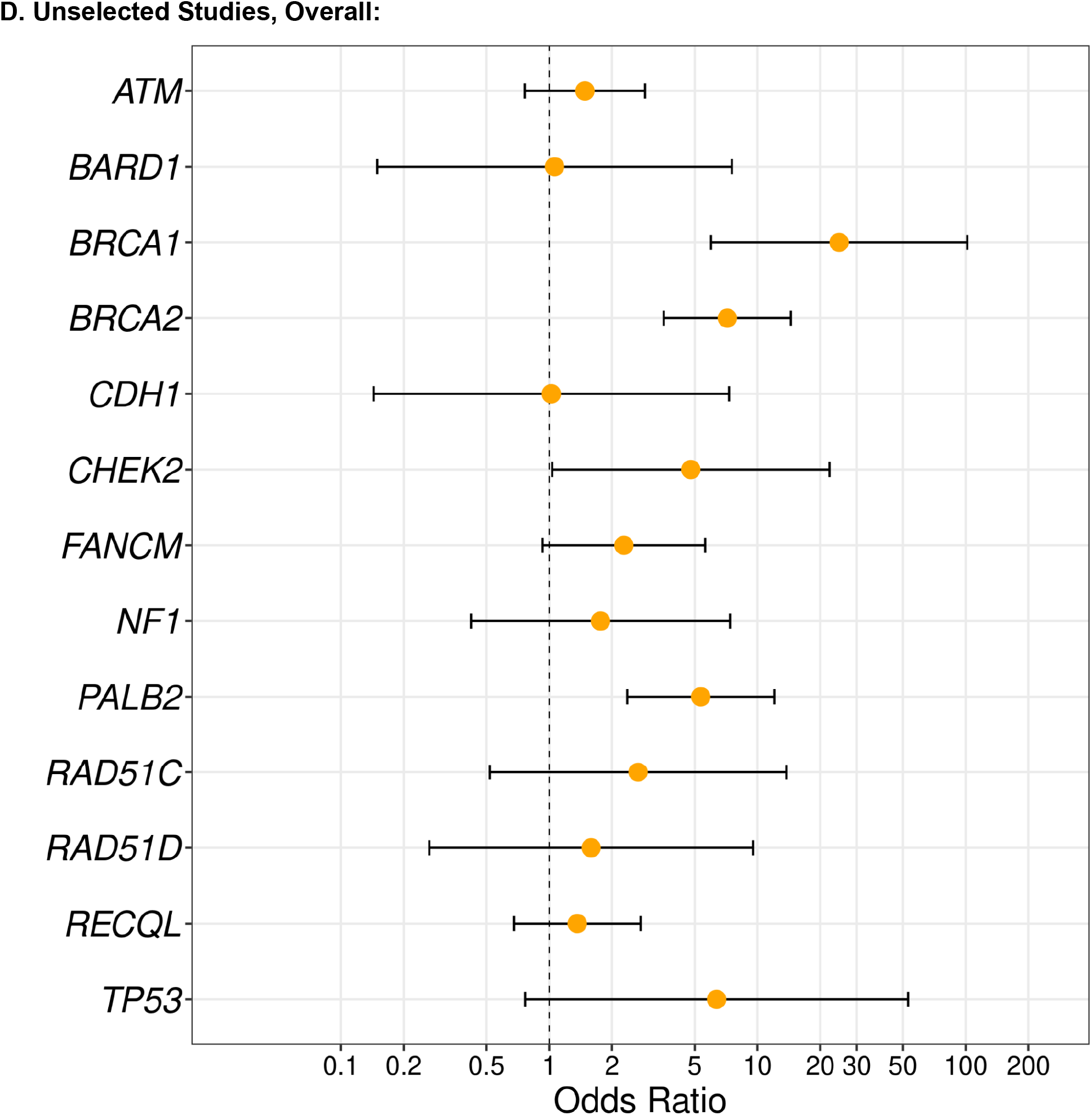
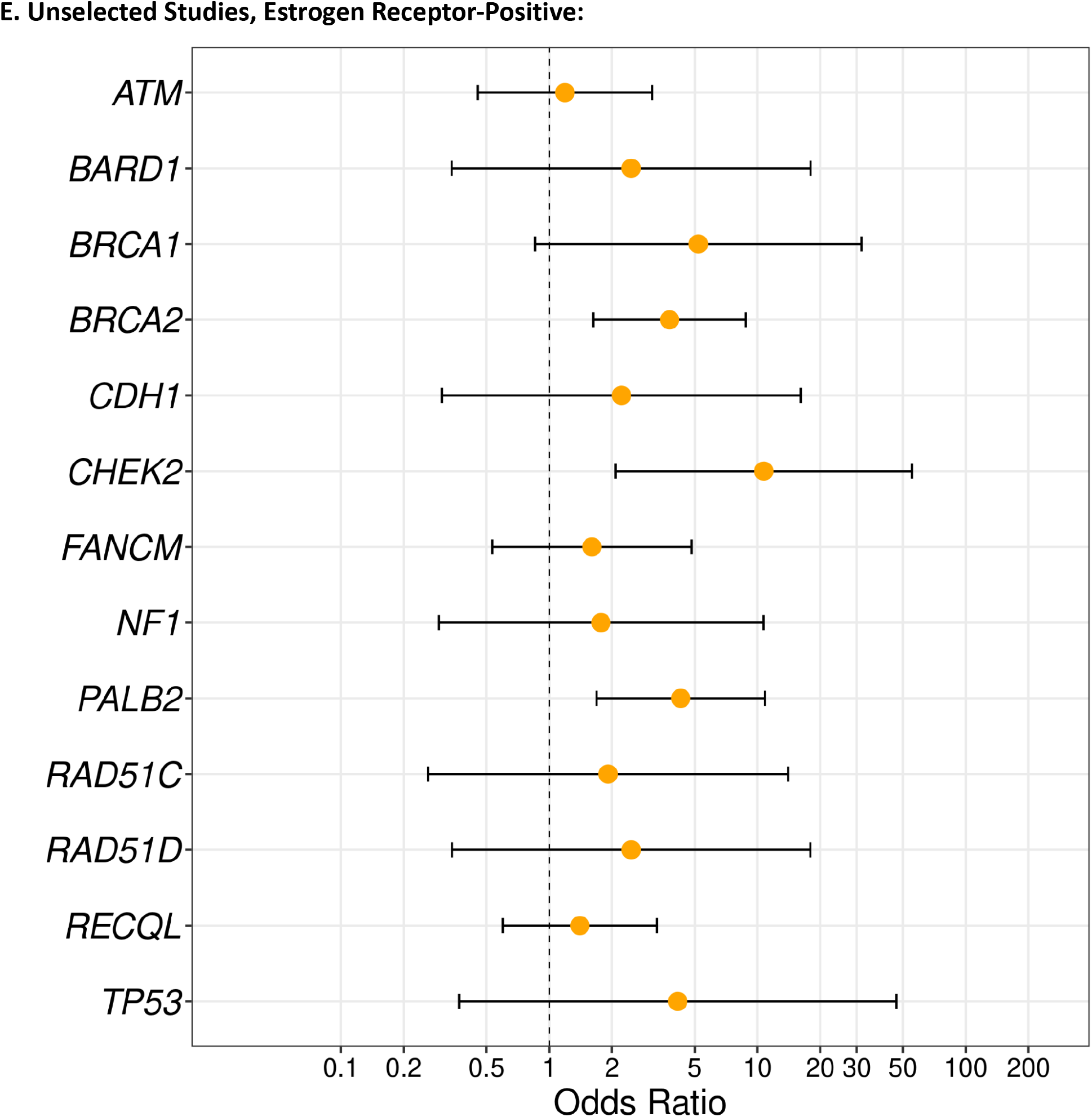
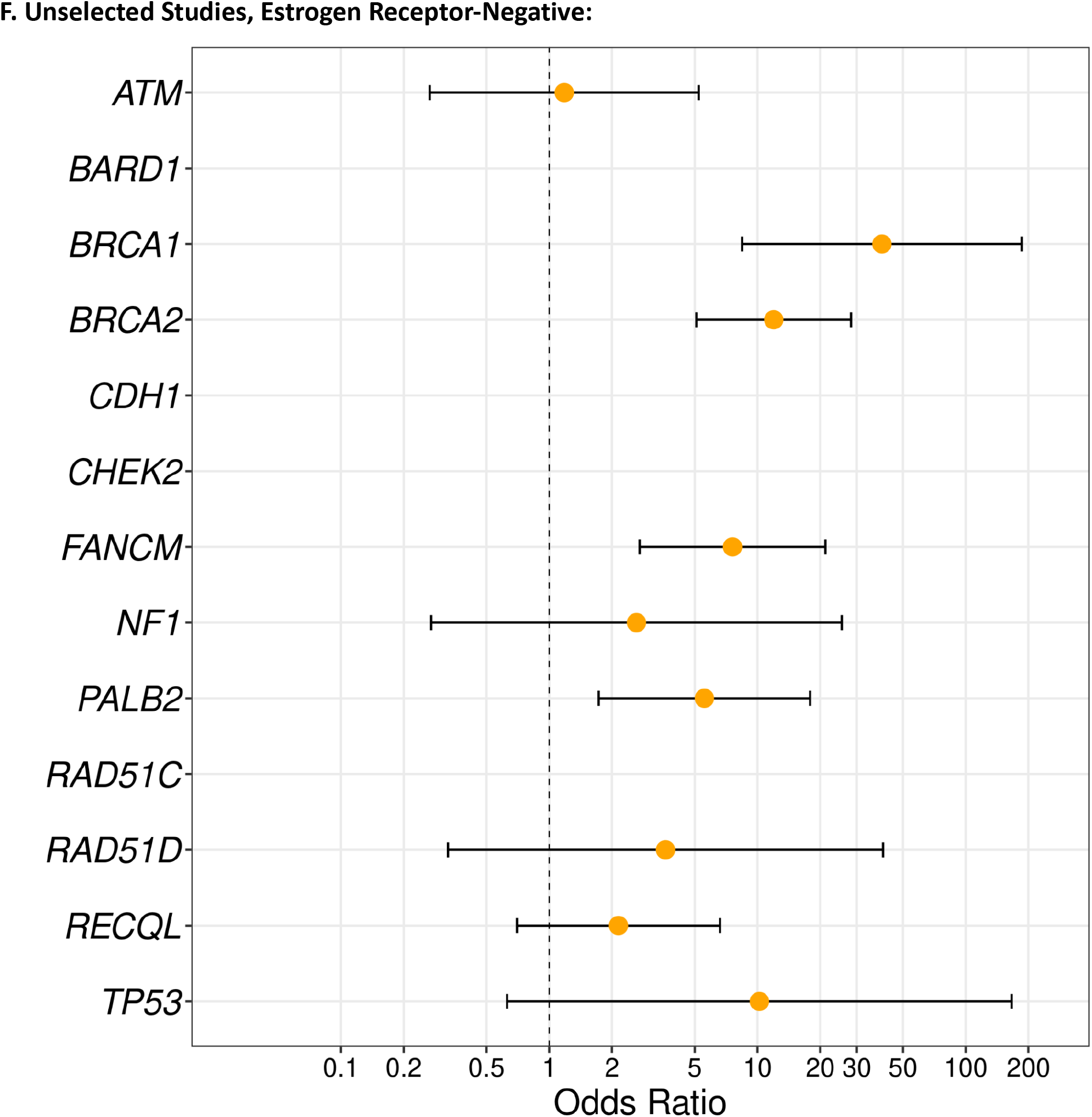
Gene-Based Odds Ratios from Joint Analysis for Breast Cancer Overall, ER-Positive, and ER-Negative Disease, in Hereditary Studies and Unselected Studies Separately. Odds ratios and confidence intervals are presented for participants in hereditary studies with overall breast cancer (Panel A), estrogen receptor (ER)-positive (Panel B), and ER-negative (Panel C) disease, and for participants in unselected studies with overall breast cancer (Panel D), estrogen receptor-positive (Panel E), and estrogen receptor-negative (Panel F) disease. The orange dot represents the point estimate and the bars represent the upper and lower bounds of the 95% confidence intervals. The X-axis describes the odds ratio on a log scale, the Y-axis represents the individual genes. Genes are listed in alphabetical order. *BRCA1* and *BRCA2* are not included for hereditary studies as participants in these studies were selected for being *BRCA1/2* negative. Participants selected for hereditary risk in the Northern California Breast Cancer Family Registry were excluded from this analysis as the selection criteria were different than those used in other studies.

**Supplementary Table 1:** Gene-Based Results for Known Genes from Joint Analysis without Exome-Wide Significance, for Breast Cancer Overall, ER-Positive, and ER-Negative Disease.

| Gene | Chr | All Studies |  |  |
| --- | --- | --- | --- | --- |
|  |  | Overall | ER-Positive | ER-Negative |
| *ATM* | 11 | 0.73 | 0.63 | 0.81 |
| *BARD1* | 2 | 0.44 | 0.17 | 0.67 |
| *CDH1* | 16 | 0.78 | 0.55 | N/A |
| *CHEK2* | 22 | 0.01 | $3.6 \times 10^{-5}$ | 0.71 |
| *NF1* | 17 | 0.62 | 0.46 | 0.37 |
| *RAD51C* | 17 | 0.39 | 0.34 | N/A |
| *RAD51D* | 17 | 0.27 | 0.27 | $1.9 \times 10^{-3}$ |
| *RECQL* | 12 | 0.52 | 0.75 | 0.40 |
| *TP53* | 17 | 0.11 | 1.00 | $1.6 \times 10^{-3}$ |

Chr=Chromosome; ER=estrogen receptor.
P-values are from gene-based SKAT-O analyses.

**Supplementary Table 2:** Gene-Based P-Values from Joint Analysis with Suggestive Significance, for Breast Cancer Overall, ER-Positive, and ER-Negative Disease.

| Gene | Chr | Overall | ER-Positive | ER-Negative |
| --- | --- | --- | --- | --- |
| *ACSM6* | 10 | 0.04 | 0.23 | 3.20E-03 |
| *BRCA1* | 17 | 2.30E-10 | 0.03 | 4.40E-16 |
| *BRCA2* | 13 | 8.40E-10 | 3.30E-04 | 8.00E-15 |
| *CCDC40* | 17 | 0.15 | 0.12 | 1.60E-03 |
| *CDHR2* | 5 | 0.13 | 0.47 | 3.00E-03 |
| *CEACAM8* | 19 | 0.22 | 0.19 | 4.90E-03 |
| *CHEK2* | 22 | 0.01 | 3.60E-05 | 0.71 |
| *DHRS4L2* | 14 | 3.60E-03 | 0.1 | 0.19 |
| *FANCG* | 9 | 0.15 | 0.29 | 3.50E-03 |
| *FANCM* | 14 | 9.80E-03 | 0.11 | 4.10E-07 |
| *FBP1* | 9 | 0.37 | N/A | 6.10E-03 |
| *FSHR* | 2 | 0.13 | 3.80E-03 | N/A |
| *GEMIN2* | 14 | 6.60E-03 | 0.07 | N/A |
| *GSTA1* | 6 | 7.90E-03 | 7.60E-03 | 0.26 |
| *LCP2* | 5 | 0.15 | 0.31 | 1.80E-03 |
| *MAPK12* | 22 | 8.30E-03 | 0.05 | 0.05 |
| *MBP* | 18 | 3.90E-04 | 7.60E-04 | 0.51 |
| *MKI67* | 10 | 0.42 | 0.57 | 3.20E-04 |
| *PALB2* | 16 | 1.80E-08 | 1.30E-05 | 5.90E-05 |
| *PRDM2* | 1 | 6.30E-03 | 0.02 | 0.57 |
| *RAD51D* | 17 | 0.27 | 0.27 | 1.90E-03 |
| *SAMD15* | 14 | 0.02 | 0.01 | 2.90E-03 |
| *SELP* | 1 | 7.10E-03 | 0.05 | 0.17 |
| *SLC26A5* | 7 | 0.06 | 0.02 | 4.40E-03 |
| *TP53* | 17 | 0.11 | 1 | 1.60E-03 |
| *TTC4* | 1 | 7.00E-03 | 6.00E-04 | 0.2 |
| *TTLL9* | 20 | 3.80E-03 | 4.40E-03 | N/A |
| *WDR93* | 15 | 0.03 | 0.56 | 1.60E-03 |
| *ZNF404* | 19 | 0.01 | 1.20E-04 | N/A |
| *ZSCAN22* | 19 | 0.03 | 4.60E-03 | 0.93 |

Chr=Chromosome; ER=estrogen receptor;
P-values are from gene-based SKAT-O analyses. Genes with $P < 0.01$ in any of the three analyses are included in the table.
\* Discovery participants were selected for being *BRCA1/2* negative (see methods), replication results are presented for *BRCA1/2*.

**Supplementary Table 3:** Gene-Based Odds Ratios and 95% Confidence Intervals from Joint Analysis for Breast Cancer Overall, ER-Positive, and ER-Negative Disease, for Genes with Suggestive Significance.

| Gene | Chromosome | Overall | ER-Positive | ER-Negative |
| --- | --- | --- | --- | --- |
| *ACSM6* | 10 | 1.81 (0.72 - 4.54) | 1.44 (0.47 - 4.39) | 4.29 (1.34 - 13.74) |
| *BRCA1\** | 17 | 24.90 (6.05 - 102.50) | 6.00 (1.08 - 33.41) | 40.73 (8.90 - 186.50) |
| *BRCA2\** | 13 | 6.96 (3.45 - 14.03) | 3.89 (1.70 - 8.89) | 10.51 (4.47 - 24.73) |
| *CCDC40* | 17 | 0.62 (0.52 - 0.74) | 0.57 (0.46 - 0.70) | 0.51 (0.36 - 0.73) |
| *CDHR2* | 5 | 2.61 (0.51 - 13.46) | 0.73 (0.07 - 8.06) | 6.52 (0.89 - 47.68) |
| *CEACAM8* | 19 | 2.61 (0.51 - 13.46) | 2.81 (0.38 - 20.86) | 7.06 (0.98 - 50.60) |
| *CHEK2* | 22 | 4.75 (1.35 - 16.69) | 9.77 (2.61 - 36.52) | NA (NA - NA) |
| *DHRS4L2* | 14 | 1.10 (0.99 - 1.23) | 1.06 (0.93 - 1.21) | 1.21 (1.00 - 1.47) |
| *FANCG* | 9 | 1.80 (0.53 - 6.16) | 0.91 (0.16 - 5.10) | 6.44 (1.60 - 25.98) |
| *FANCM* | 14 | 2.68 (1.29 - 5.54) | 2.20 (0.94 - 5.16) | 6.69 (2.86 - 15.65) |
| *FBP1* | 9 | 1.42 (0.24 - 8.49) | N/A | 4.93 (0.69 - 35.41) |
| *FSHR* | 2 | 5.16 (0.60 - 44.26) | 12.04 (1.35 - 107.49) | N/A |
| *GEMIN2* | 14 | 0.14 (0.02 - 1.10) | 0.35 (0.04 - 2.83) | N/A |
| *GSTA1* | 6 | 2.87 (1.28 - 6.43) | 3.20 (1.32 - 7.80) | 2.13 (0.56 - 8.14) |
| *LCP2* | 5 | 5.01 (0.58 - 42.93) | 4.30 (0.38 - 49.03) | 12.46 (1.11 - 140.47) |
| *MAPK12* | 22 | 0.48 (0.28 - 0.80) | 0.42 (0.20 - 0.90) | 0.29 (0.07 - 1.19) |
| *MBP* | 18 | 4.05 (1.66 - 9.89) | 4.31 (1.64 - 11.36) | 1.80 (0.36 - 9.09) |
| *MKI67* | 10 | 1.24 (0.53 - 2.87) | 1.49 (0.56 - 3.99) | 2.44 (0.76 - 7.87) |
| *PALB2* | 16 | 6.47 (3.19 - 13.11) | 5.11 (2.33 - 11.19) | 6.43 (2.51 - 16.48) |
| *PRDM2* | 1 | 1.95 (1.04 - 3.65) | 2.27 (1.12 - 4.61) | 1.06 (0.30 - 3.69) |
| *RAD51D* | 17 | 2.00 (0.37 - 10.91) | 2.08 (0.29 - 15.18) | 5.35 (0.75 - 38.18) |
| *SAMD15* | 14 | 1.45 (0.73 - 2.88) | 1.66 (0.72 - 3.82) | 3.12 (1.24 - 7.87) |
| *SELP* | 1 | 0.23 (0.08 - 0.69) | 0.23 (0.05 - 0.98) | N/A |
| *SLC26A5* | 7 | 2.06 (1.21 - 3.52) | 1.94 (1.04 - 3.59) | 2.04 (0.89 - 4.67) |
| *TP53* | 17 | 7.22 (0.89 - 58.53) | 3.42 (0.30 - 38.46) | 15.47 (1.39 - 172.75) |
| *TTC4* | 1 | 2.94 (0.80 - 10.87) | 4.98 (1.30 - 18.99) | 1.56 (0.16 - 15.24) |
| *TTLL9* | 20 | 0.26 (0.10 - 0.70) | 0.09 (0.01 - 0.68) | N/A |
| *WDR93* | 15 | 2.62 (0.82 - 8.35) | 1.21 (0.22 - 6.68) | 4.84 (1.07 - 21.91) |
| *ZNF404* | 19 | 4.78 (1.36 - 16.80) | 7.35 (1.94 - 27.90) | N/A |
| *ZSCAN22* | 19 | 1.90 (0.95 - 3.82) | 2.50 (1.18 - 5.31) | 0.94 (0.21 - 4.22) |

ER=estrogen receptor
\* Discovery participants were selected for being *BRCA1/2* negative (see methods), replication results are presented for *BRCA1/2*.

**Supplementary Table 4:** Gene-Based P-Values from Joint Analysis Including Missense Variants with Suggestive Significance for Breast Cancer Overall, ER-Positive, and ER-Negative Disease.

| Gene | Chr | Overall | ER-Positive | ER-Negative |
| --- | --- | --- | --- | --- |
| *ACSM6* | 10 | 0.04 | 0.23 | 3.20E-03 |
| *ATR* | 3 | 0.02 | 9.20E-04 | 0.29 |
| *BRCA1* | 17 | 2.30E-10 | 0.03 | 4.40E-16 |
| *BRCA2* | 13 | 6.70E-10 | 2.30E-04 | 1.30E-14 |
| *CASP8AP2* | 6 | 2.90E-03 | 0.58 | 0.13 |
| *CCDC40* | 17 | 0.1 | 0.12 | 1.20E-04 |
| *CDHR2* | 5 | 0.13 | 0.47 | 3.00E-03 |
| *CEACAM8* | 19 | 0.22 | 0.19 | 4.90E-03 |
| *CHEK2* | 22 | 4.10E-03 | 1.00E-04 | 0.4 |
| *DDX56* | 7 | 0.09 | 6.00E-03 | N/A |
| *DHRS4L2* | 14 | 3.60E-03 | 0.1 | 0.19 |
| *DSTYK* | 1 | 8.70E-03 | 0.13 | N/A |
| *FANCG* | 9 | 0.15 | 0.35 | 8.60E-03 |
| *FANCM* | 14 | 0.04 | 0.2 | 2.90E-06 |
| *FAT3* | 11 | 0.29 | 0.49 | 6.60E-03 |
| *GEMIN2* | 14 | 7.10E-03 | 0.07 | 0.25 |
| *GSTA1* | 6 | 7.90E-03 | 7.60E-03 | 0.26 |
| *LCP2* | 5 | 0.15 | 0.31 | 1.80E-03 |
| *MBP* | 18 | 3.90E-04 | 7.60E-04 | 0.51 |
| *MKI67* | 10 | 0.42 | 0.57 | 3.20E-04 |
| *MSH6* | 2 | 4.20E-03 | 3.90E-03 | 0.04 |
| *NDOR1* | 9 | 4.70E-03 | 3.10E-03 | 0.07 |
| *PALB2* | 16 | 1.80E-08 | 1.30E-05 | 5.90E-05 |
| *PCDHGC5* | 5 | 0.21 | 1.0 | 6.50E-03 |
| *PRDM2* | 1 | 4.60E-03 | 0.02 | 0.85 |
| *PREX2* | 8 | 0.25 | 0.44 | 8.60E-03 |
| *RAD51D* | 17 | 0.44 | 0.17 | 5.40E-03 |
| *SAMD15* | 14 | 0.02 | 0.01 | 2.90E-03 |
| *SDK2* | 17 | 0.02 | 9.60E-03 | 3.00E-03 |
| *SERINC3* | 20 | 0.37 | 1.0 | 1.50E-03 |
| *SLC26A5* | 7 | 0.03 | 0.02 | 4.40E-03 |
| *TTC4* | 1 | 7.00E-03 | 6.00E-04 | 0.2 |
| *TTLL9* | 20 | 6.00E-03 | 4.30E-03 | 0.1 |
| *WDR93* | 15 | 0.03 | 0.56 | 1.60E-03 |
| *ZNF404* | 19 | 0.01 | 1.20E-04 | N/A |

**Supplementary Table 5:** Gene-Based Odds Ratios and 95% Confidence Intervals from Joint Analysis Including Missense Variants for Breast Cancer Overall, ER-Positive, and ER-Negative Disease, for Genes with Suggestive Significance.

| Gene | Chromosome | Overall | ER-Positive | ER-Negative |
| --- | --- | --- | --- | --- |
| *ACSM6* | 10 | 1.81 (0.72 - 4.54) | 1.44 (0.47 - 4.39) | 4.29 (1.34 - 13.74) |
| *ATR* | 3 | 2.58 (1.14 - 5.87) | 3.84 (1.56 - 9.45) | 1.51 (0.32 - 7.20) |
| *BRCA1* | 17 | 24.90 (6.05 - 102.50) | 6.00 (1.08 - 33.41) | 40.73 (8.90 - 186.50) |
| *BRCA2* | 13 | 6.47 (3.32 - 12.63) | 3.84 (1.74 - 8.46) | 9.53 (4.15 - 21.85) |
| *CASP8AP2* | 6 | 1.15 (1.05 - 1.27) | 1.02 (0.90 - 1.15) | 0.88 (0.73 - 1.07) |
| *CCDC40* | 17 | 0.62 (0.53 - 0.74) | 0.57 (0.46 - 0.70) | 0.53 (0.37 - 0.75) |
| *CDHR2* | 5 | 3.12 (0.63 - 15.48) | 1.63 (0.23 - 11.69) | 6.52 (0.89 - 47.68) |
| *CEACAM8* | 19 | 2.61 (0.51 - 13.46) | 2.81 (0.38 - 20.86) | 7.06 (0.98 - 50.60) |
| *CHEK2* | 22 | 4.01 (1.50 - 10.77) | 5.98 (2.04 - 17.50) | 1.31 (0.15 - 11.44) |
| *DDX56* | 7 | 6.20 (0.75 - 51.53) | 11.81 (1.34 - 104.03) | N/A |
| *DHRS4L2* | 14 | 1.11 (1.00 - 1.23) | 1.06 (0.93 - 1.21) | 1.21 (1.00 - 1.47) |
| *DSTYK* | 1 | 0.09 (0.01 - 0.70) | 0.18 (0.02 - 1.37) | N/A |
| *FANCG* | 9 | 1.43 (0.45 - 4.51) | 0.72 (0.14 - 3.80) | 5.01 (1.33 - 18.83) |
| *FANCM* | 14 | 2.01 (1.08 - 3.74) | 1.79 (0.85 - 3.77) | 4.61 (2.13 - 9.96) |
| *FAT3* | 11 | 1.58 (0.71 - 3.49) | 1.46 (0.54 - 3.89) | 3.60 (1.34 - 9.62) |
| *GEMIN2* | 14 | 0.27 (0.06 - 1.27) | 0.35 (0.04 - 2.83) | 0.86 (0.11 - 6.88) |
| *GSTA1* | 6 | 2.87 (1.28 - 6.43) | 3.20 (1.32 - 7.80) | 2.13 (0.56 - 8.14) |
| *LCP2* | 5 | 5.01 (0.58 - 42.93) | 4.30 (0.38 - 49.03) | 12.46 (1.11 - 140.47) |
| *MBP* | 18 | 4.05 (1.66 - 9.89) | 4.31 (1.64 - 11.36) | 1.80 (0.36 - 9.09) |
| *MKI67* | 10 | 1.24 (0.53 - 2.87) | 1.49 (0.56 - 3.99) | 2.44 (0.76 - 7.87) |
| *MSH6* | 2 | 0.90 (0.81 - 0.99) | 1.34 (1.19 - 1.51) | 1.61 (1.34 - 1.92) |
| *NDOR1* | 9 | 3.09 (1.51 - 6.33) | 3.61 (1.63 - 7.97) | 3.23 (1.09 - 9.51) |
| *PALB2* | 16 | 6.47 (3.19 - 13.11) | 5.11 (2.33 - 11.19) | 6.43 (2.51 - 16.48) |
| *PCDHGC5* | 5 | 1.07 (0.98 - 1.16) | 0.88 (0.79 - 0.98) | 0.86 (0.73 - 1.02) |
| *PRDM2* | 1 | 1.17 (0.71 - 1.94) | 1.33 (0.73 - 2.43) | 1.05 (0.40 - 2.74) |
| *PREX2* | 8 | 1.07 (0.50 - 2.29) | 0.43 (0.12 - 1.53) | 2.68 (1.01 - 7.15) |
| *RAD51D* | 17 | 1.53 (0.43 - 5.45) | 2.31 (0.57 - 9.38) | 2.98 (0.54 - 16.44) |
| *SAMD15* | 14 | 1.45 (0.73 - 2.88) | 1.66 (0.72 - 3.82) | 3.12 (1.24 - 7.87) |
| *SDK2* | 17 | 4.20 (0.89 - 19.78) | 5.14 (0.93 - 28.36) | 6.71 (0.94 - 48.04) |
| *SERINC3* | 20 | 1.40 (0.44 - 4.41) | 0.81 (0.15 - 4.34) | 4.67 (1.23 - 17.76) |
| *SLC26A5* | 7 | 2.12 (1.25 - 3.62) | 1.94 (1.04 - 3.59) | 2.04 (0.89 - 4.67) |
| *TTC4* | 1 | 2.94 (0.80 - 10.87) | 4.98 (1.30 - 18.99) | 1.56 (0.16 - 15.24) |
| *TTLL9* | 20 | 0.43 (0.21 - 0.91) | 0.15 (0.04 - 0.64) | 0.48 (0.11 - 2.03) |
| *WDR93* | 15 | 2.84 (0.90 - 8.94) | 1.65 (0.36 - 7.53) | 4.84 (1.07 - 21.91) |
| *ZNF404* | 19 | 4.78 (1.36 - 16.80) | 7.35 (1.94 - 27.90) | N/A |

ER=estrogen receptor
\* Discovery participants were selected for being *BRCA1/2* negative (see methods), replication results are presented for *BRCA1/2*.

## Supplementary Methods

**Genes Selected for Replication Based on Discovery Findings**

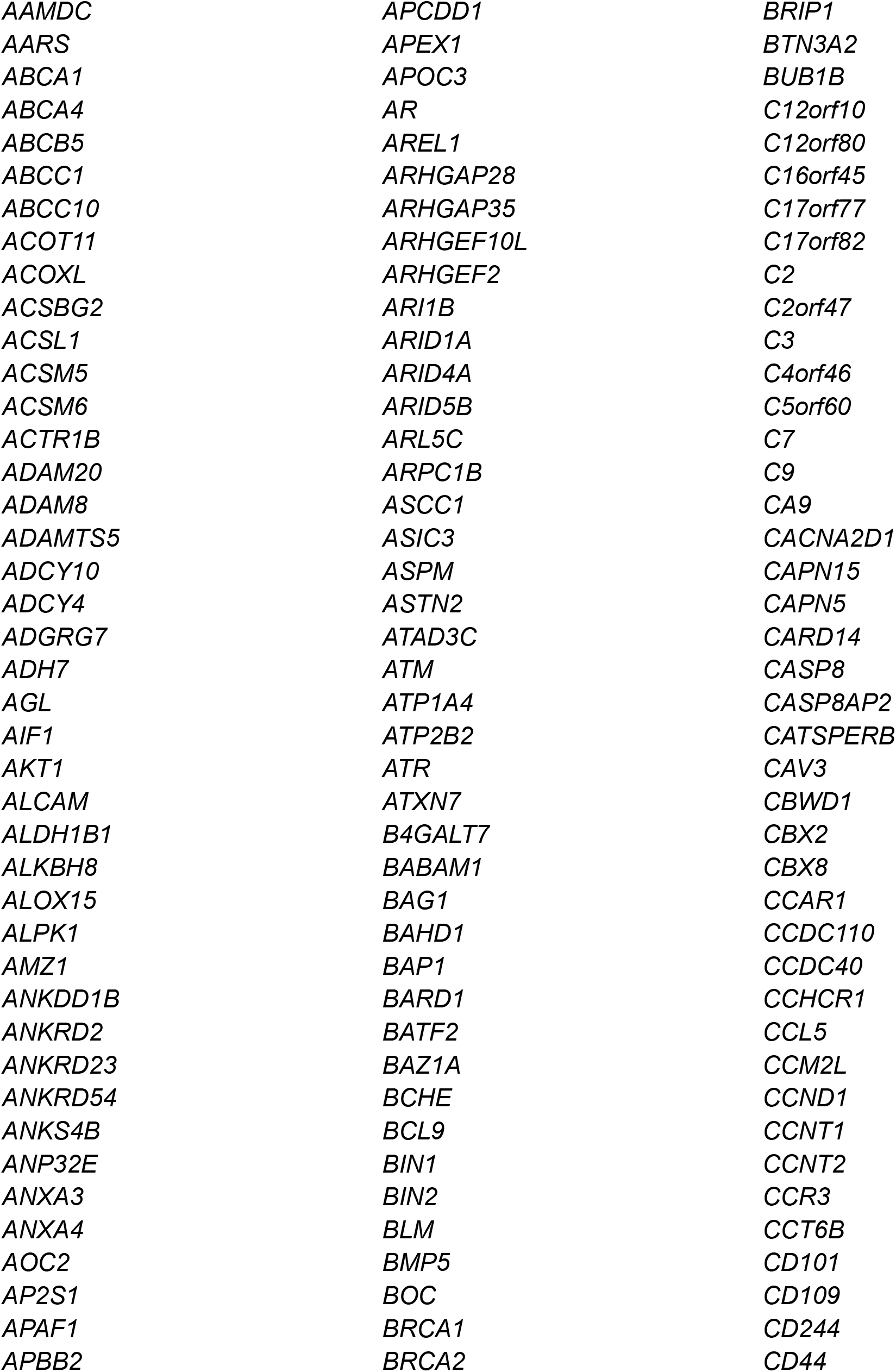
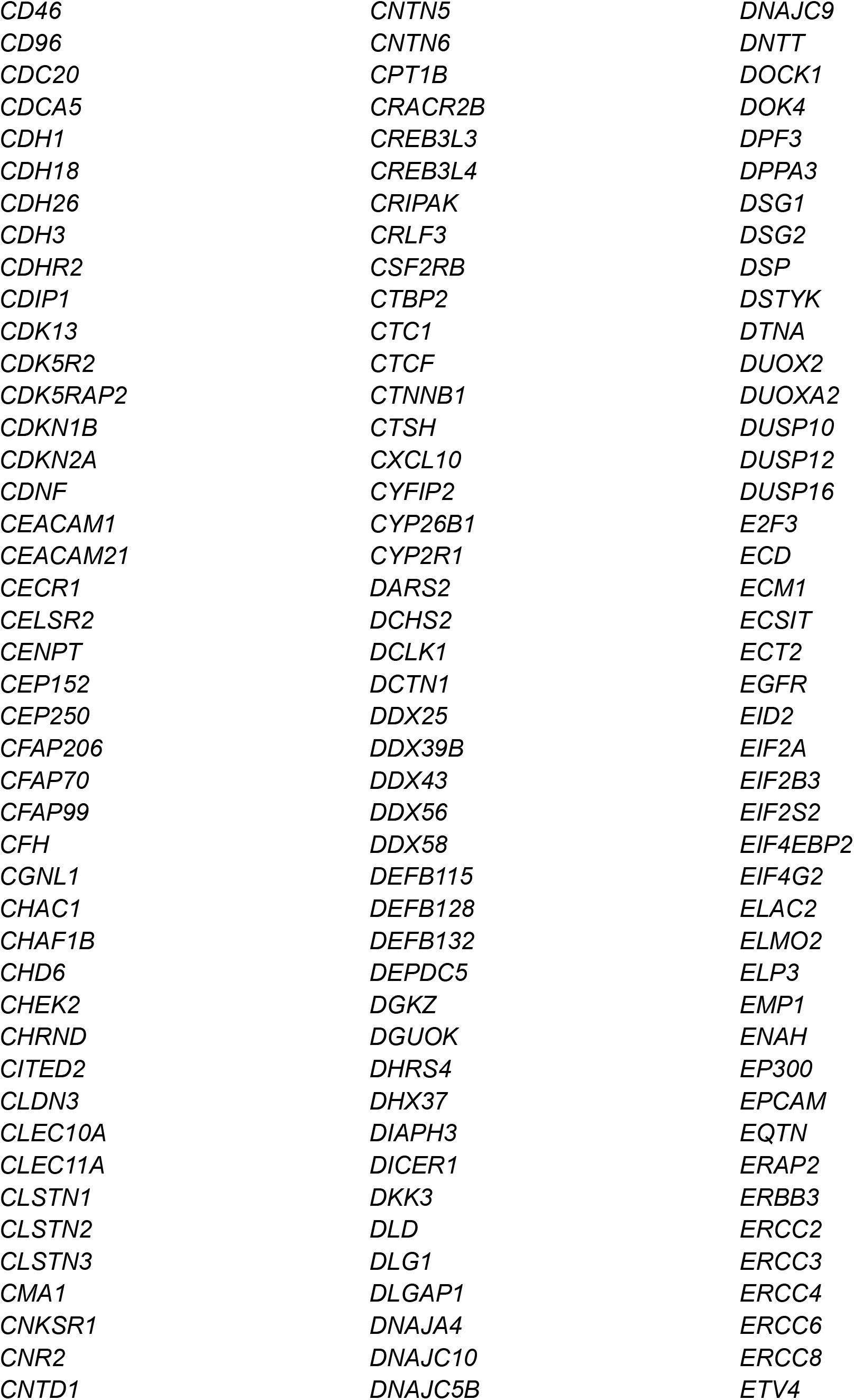
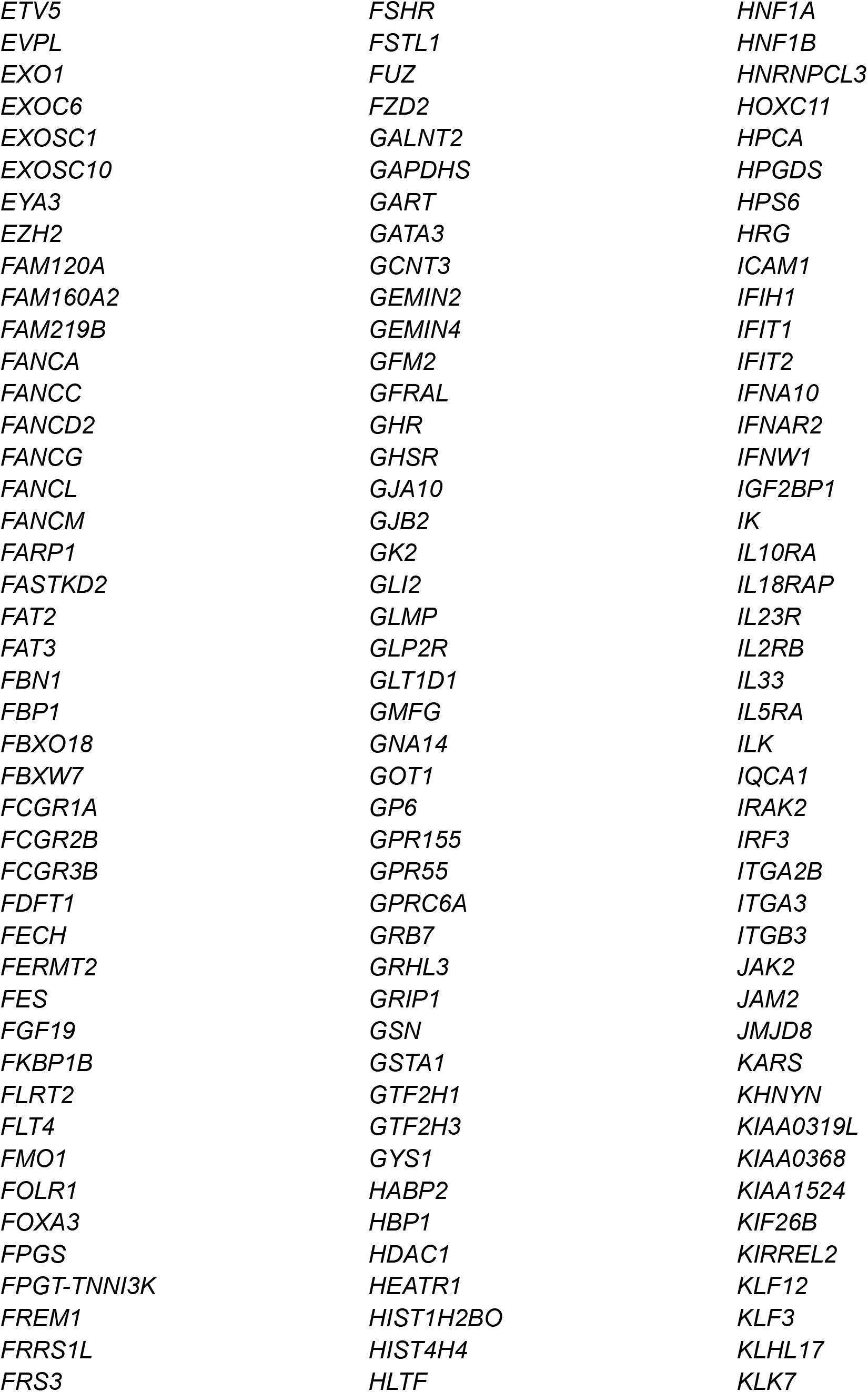
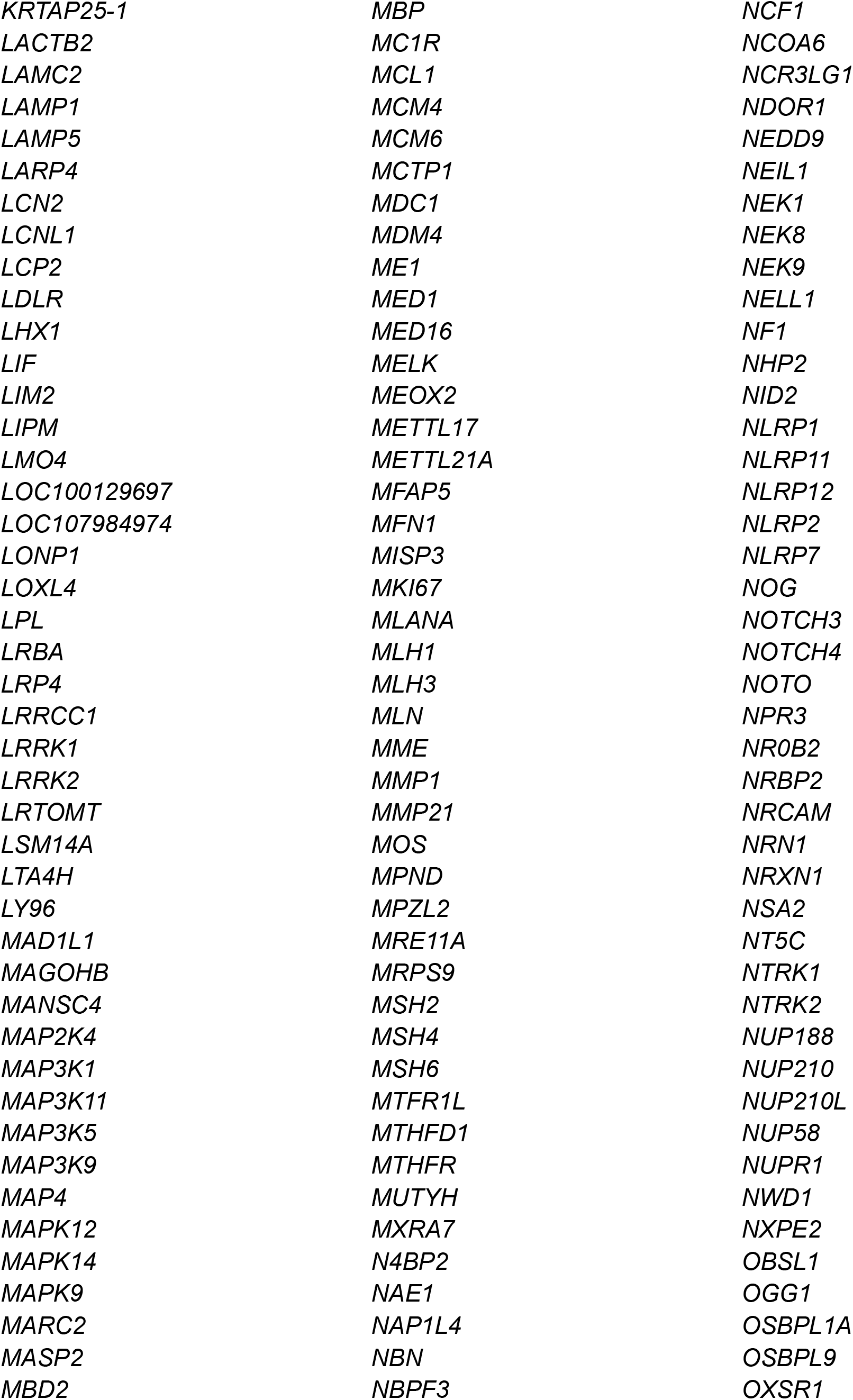
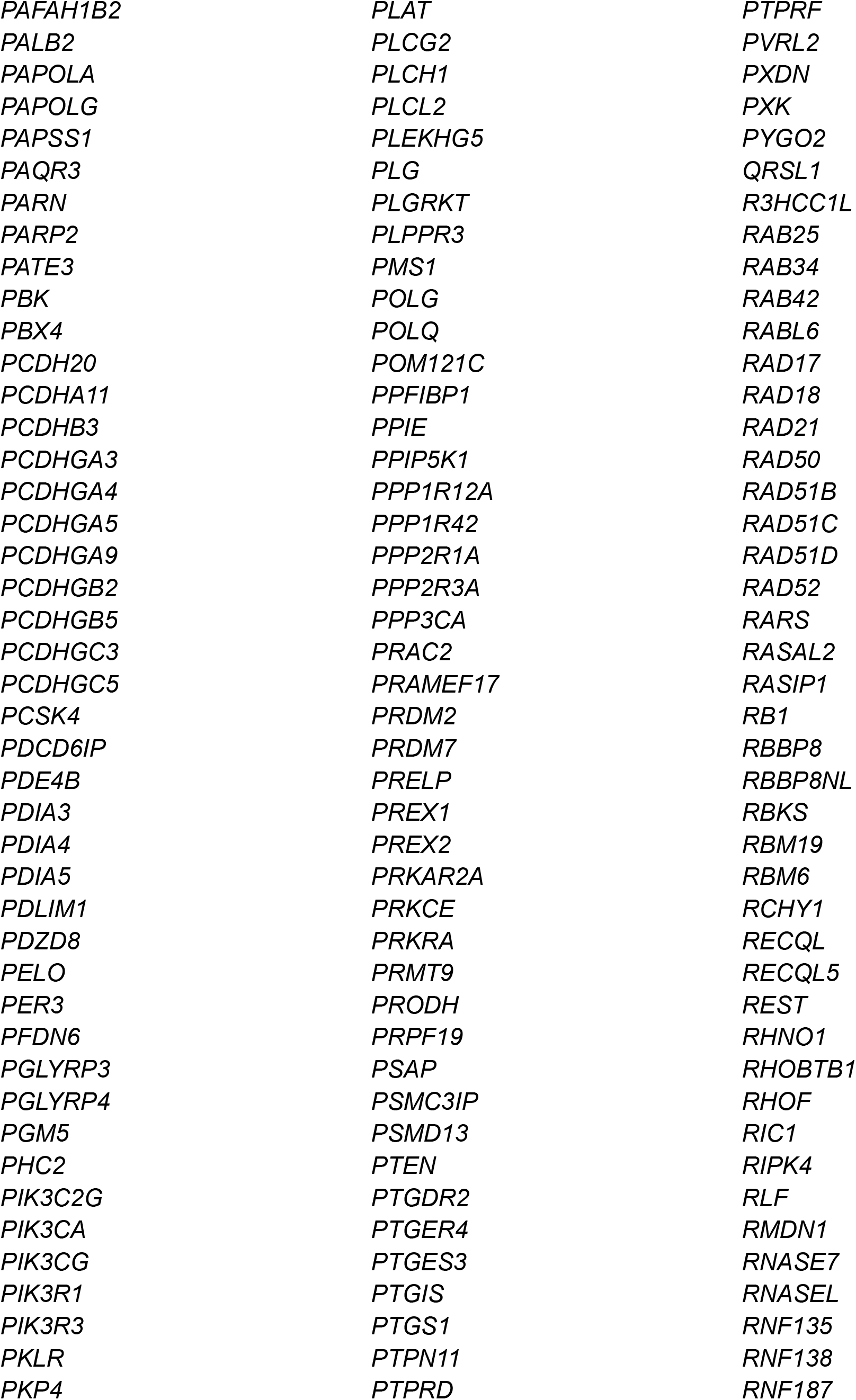
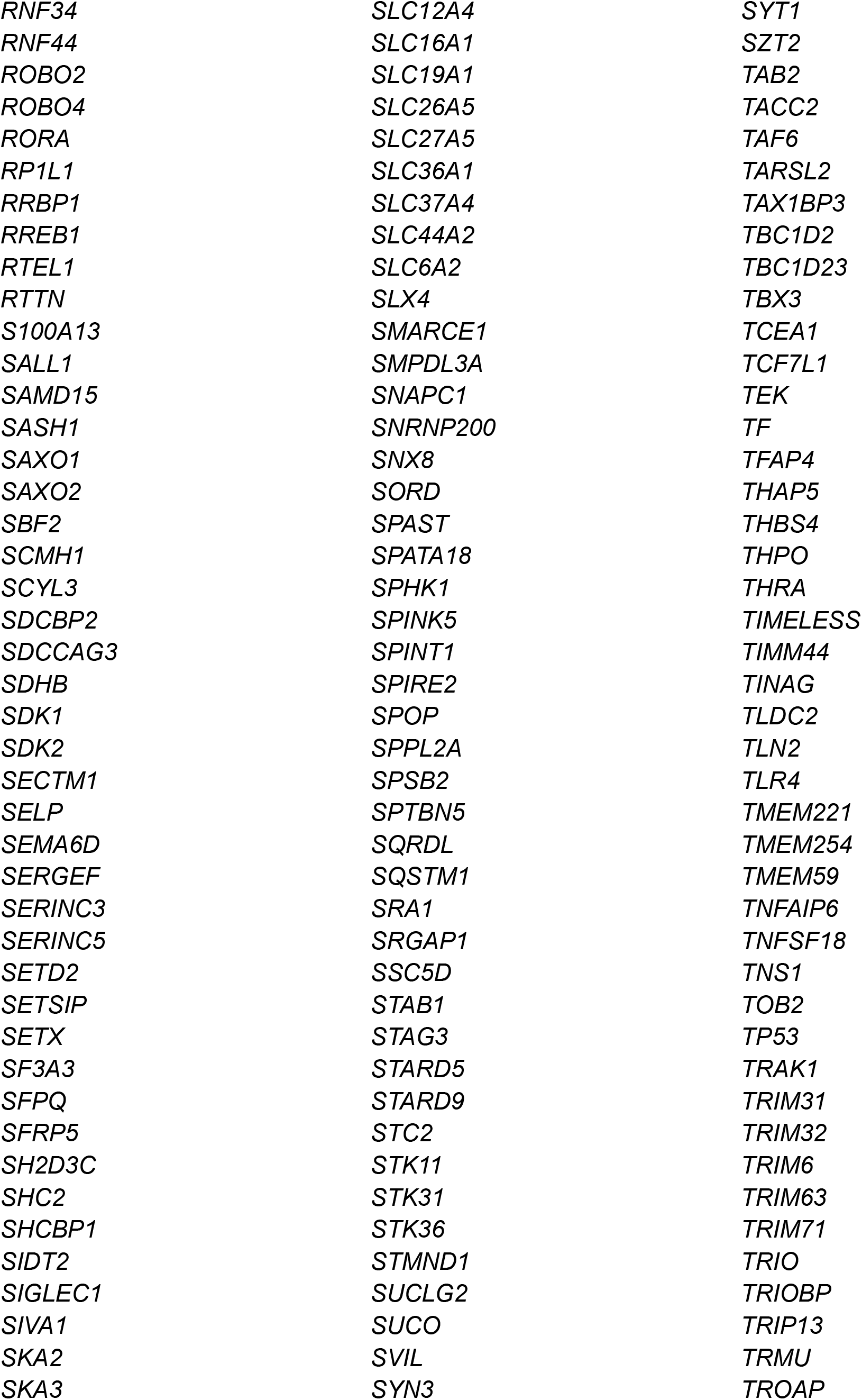
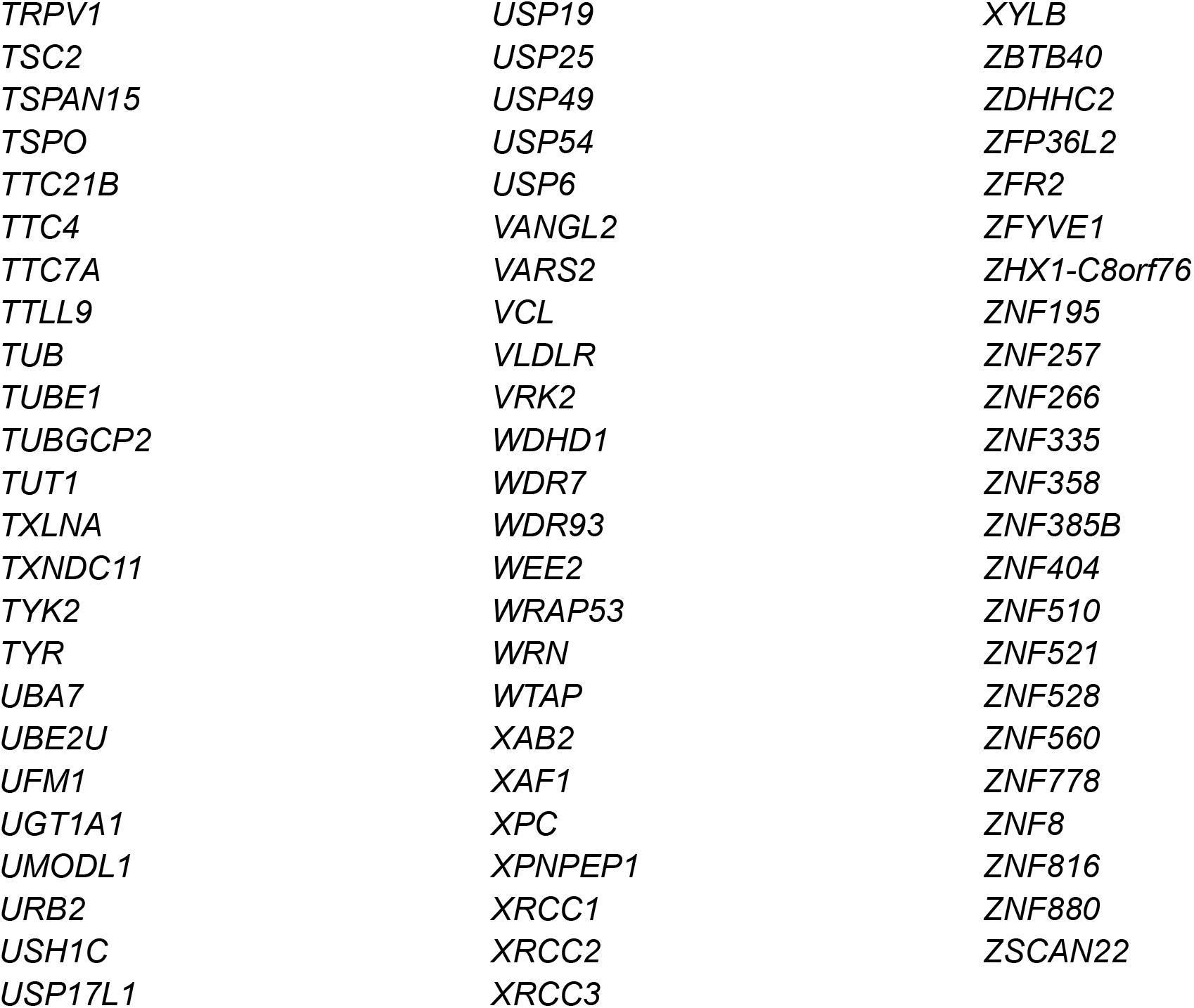

